# GWAS and meta-analysis identifies multiple new genetic mechanisms underlying severe Covid-19

**DOI:** 10.1101/2022.03.07.22271833

**Authors:** Erola Pairo-Castineira, Konrad Rawlik, Lucija Klaric, Athanasios Kousathanas, Anne Richmond, Jonathan Millar, Clark D Russell, Tomas Malinauskas, Ryan Thwaites, Alex Stuckey, Christopher A Odhams, Susan Walker, Fiona Griffiths, Wilna Oosthuyzen, Kirstie Morrice, Sean Keating, Alistair Nichol, Malcolm G Semple, Julian Knight, Manu Shankar-Hari, Charlotte Summers, Charles Hinds, Peter Horby, Lowell Ling, Danny McAuley, Hugh Montgomery, Peter J.M. Openshaw, Timothy Walsh, Albert Tenesa, GenOMICC Investigators, SCOURGE Consortium, ISARIC4C Investigators, 23andMe, Richard H Scott, Mark J Caulfield, Loukas Moutsianas, Chris P Ponting, James F Wilson, Veronique Vitart, Alexandre C Pereira, Andre Luchessi, Esteban Parra, Raquel Cruz-Guerrero, Angel Carracedo, Angie Fawkes, Lee Murphy, Kathy Rowan, Andy Law, Sara Clohisey Hendry, J. Kenneth Baillie

## Abstract

Pulmonary inflammation drives critical illness in Covid-19, ^1;2^ creating a clinically homogeneous extreme phenotype, which we have previously shown to be highly efficient for discovery of genetic associations. ^3;4^ Despite the advanced stage of illness, we have found that immunomodulatory therapies have strong beneficial effects in this group. ^1;5^ Further genetic discoveries may identify additional therapeutic targets to modulate severe disease. ^6^ In this new data release from the GenOMICC (Genetics Of Mortality in Critical Care) study we include new microarray genotyping data from additional critically-ill cases in the UK and Brazil, together with cohorts of severe Covid-19 from the ISARIC4C ^7^ and SCOURGE ^8^ studies, and meta-analysis with previously-reported data. We find an additional 14 new genetic associations. Many are in potentially druggable targets, in inflammatory signalling (JAK1, PDE4A), monocyte-macrophage differentiation (CSF2), immunometabolism (SLC2A5, AK5), and host factors required for viral entry and replication (TMPRSS2, RAB2A). As with our previous work, these results provide tractable therapeutic targets for modulation of harmful host-mediated inflammation in Covid-19.

## Introduction

Critical illness in Covid-19 is a relatively narrow clinical phenotype characterised by hypoxaemic respiratory failure ^9^ in which lung injury is, at least in part, caused by host-mediated inflammation. ^1;2^ The international GenOMICC (Genetics of Mortality in Critical Care) consortium has shown that critical illness is a highly efficient phenotype for discovery of genetic variants associated with Covid-19, ^3;4^ contributing the largest signal to recent meta-analyses ^10^ despite only including a relatively small number of patients. This is consistent with our previous predictions for other respiratory viral illnesses. ^11;12^ New discoveries in the first GenOMICC study (implicating OAS1, TYK2, IFNAR2, and DPP9), ^3^ completed only 5 months after the first patient presented to a participating intensive care unit, led to new understanding of the host:pathogen interaction, ^3;13;14^ and, in part, led to the discovery of a new effective treatment for severe Covid-19. ^15^

Here, we provide an updated analysis of the international GenOMICC study comprising a combination of microarray genotype data from 11,325 critically ill cases in the UK (9,279 cases) and Brazil (2,186), combined with other studies recruiting hospitalised patients with a strong focus on severe and critical disease: ISARIC4C (655 cases) and SCOURGE consortium (5,934 cases)(Table 1). We put these new results in context by completing a meta-analysis of the new GenOMICC GWAS results with previously-published data.

**Table 1:** Description of cohorts included in the critical meta-analysis with median age and percentage of females for the cases. NA: information not available for this cohort. * mean was provided rather than the median. ** data was available for the hospitalisation cohort but not for the critically-ill patients.

| Study | Dataset | Ancestry | Case definition | Control definition | Median age(IQR) | Female(perc) | N cases | Ncontrols |
| --- | --- | --- | --- | --- | --- | --- | --- | --- |
| GenOMICC | Whole-genome Sequence | EUR | Critical care | General population and mild cases | 60(11.87) | 32.17% | 5989 | 42981 |
| GenOMICC | Whole-genome Sequence | EAS | Critical care | General population and mild cases | 54(11.27) | 40.87% | 274 | 366 |
| GenOMICC | Whole-genome Sequence | SAS | Critical care | General population and mild cases | 57(13.22) | 25.63% | 788 | 3793 |
| GenOMICC | Whole-genome Sequence | AFR | Critical care | General population and mild cases | 57(13.06) | 35% | 440 | 1350 |
| GenOMICC | Genotype | EUR | Critical care | General population | 58(13.49) | 32.4% | 1347 | 6735 |
| GenOMICC | Genotype | SAS | Critical care | General population | 58.5(16.10) | 28.5% | 200 | 1000 |
| GenOMICC | Genotype | AFR | Critical care | General population | 55(16.65) | 44.5% | 101 | 505 |
| ISARIC4C | Genotype | EUR | ICU/CPAP/NIV | General population | 59(12.75) | 30% | 140 | 700 |
| HGIv6 | Genotype | ALL | Critical (A2) | General population | NA | NA | 8779 | 1001875 |
| SCOURGE | Genotype | EUR | SCOURGE severe 3 -4 | General population | 69.13(12.93)* | 27.5% | 3502 | 5455 |
| 23 and me | Genotype | EUR | Respiratory Support | General population | 53(17.5)** | 50.7% ** | 495 | 680440 |

## Results

The design of the GenOMICC study has been previously described. ^3;4^ Briefly, patients with confirmed Covid-19 requiring continuous cardiorespiratory monitoring or organ support (a generalisable definition for admission to a critical care area) were recruited in 2020-21. Using participants recruited and genotyped in GenOMICC and ISARIC4C since the first reported GenOMICC GWAS, ^3^ we performed ancestry-specific GWAS largely following the methods we previously described ^3;4^ (see Material and Methods, and Supplementary Material). Using the results of these GWAS, previously reported results obtained using GenOMICC participants with whole genome sequencing data ^4^, and data from GenOMICC Brazil, we performed trans-ancestry and -platform meta analyses, within the GenOMICC study, for a critically ill Covid-19 phenotype and a hospitalised Covid-19 phenotype (see Material and Methods). Results of these GenOMICC-only meta analyses are illustrated in the lower parts of Figure 1 and Supplementary Figure 3 for the critically ill and hospitalised phenotypes respectively.

**Figure 1:**
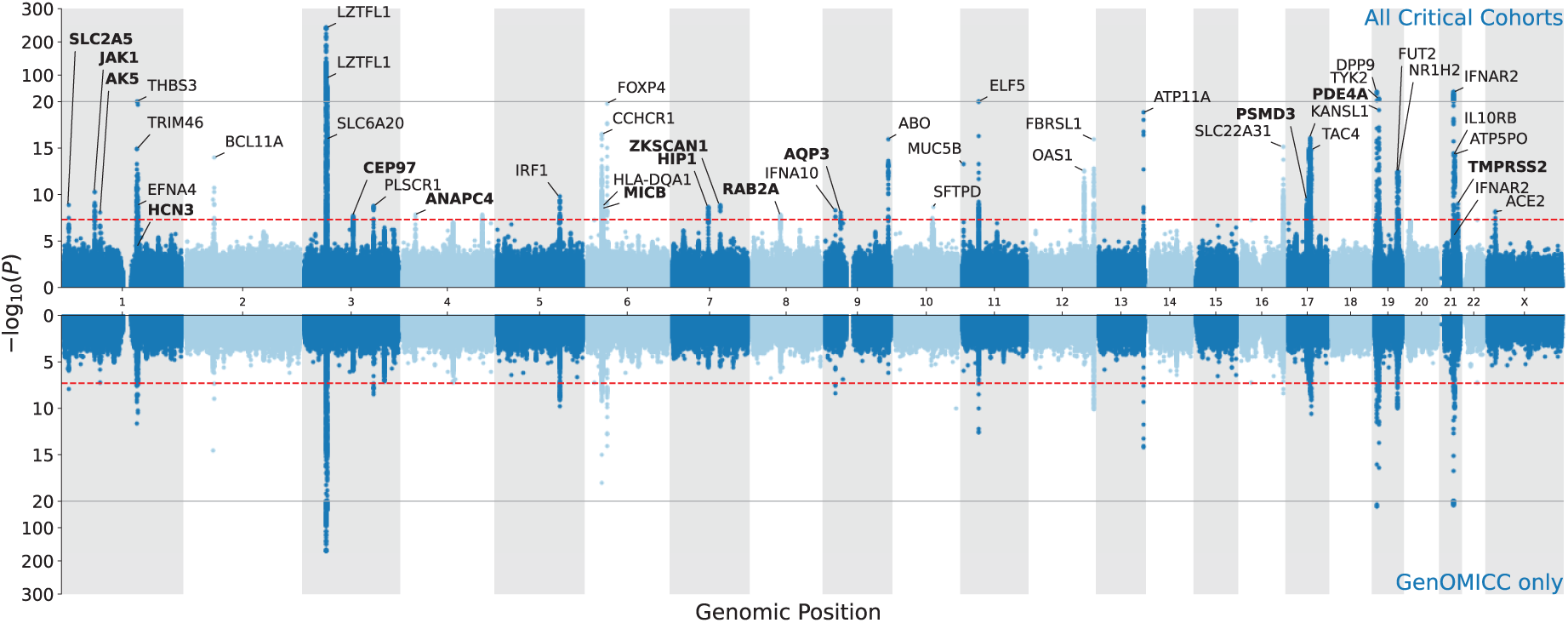
Miami plot showing meta-analysis results obtained using all critical phenotype cohorts (top) and using GenOMICC data only (bottom). Independent lead variants in the all critical cohorts analysis are annotated with nearby or plausible genes, with new associations from the present study in bold.

In order to put these new findings in the context of existing knowledge, we performed comprehensive meta analyses, drawing on further GWAS results for the two phenotypes of interest, including data kindly shared by the SCOURGE consortium and published data from the the Covid-19 Human Genetics Initiative (HGIv6, 2021). Characteristics of contributing studies to the meta analyses are summarised in Table 1 and Supplementary Table 1 for the critically ill and hospitalised phenotypes, with further details on each study provided in the Supplementary Material. Since leave-one-out analyses from the HGIv6 release were not available, we used a mathematical subtraction approach, as in our previous work, ^4^,to remove the GenOMICC signals from HGIv6, yielding an independent data set. This was essential because previous GenOMICC data contribute 20 to 50% of each genetic association signal in the HGI analyses. The results of these All Critical Cohorts and All Hospitalised Cohorts meta analyses are illustrated in Figure 1 and Supplementary Figure 3. Independent lead variants for the critically ill phenotype are summarised in Table 2 with more details provided in Supplementary Table 2, while Supplementary Table 4 contains details of lead variants from the analysis of the hospitalised phenotype.

**Table 2:**
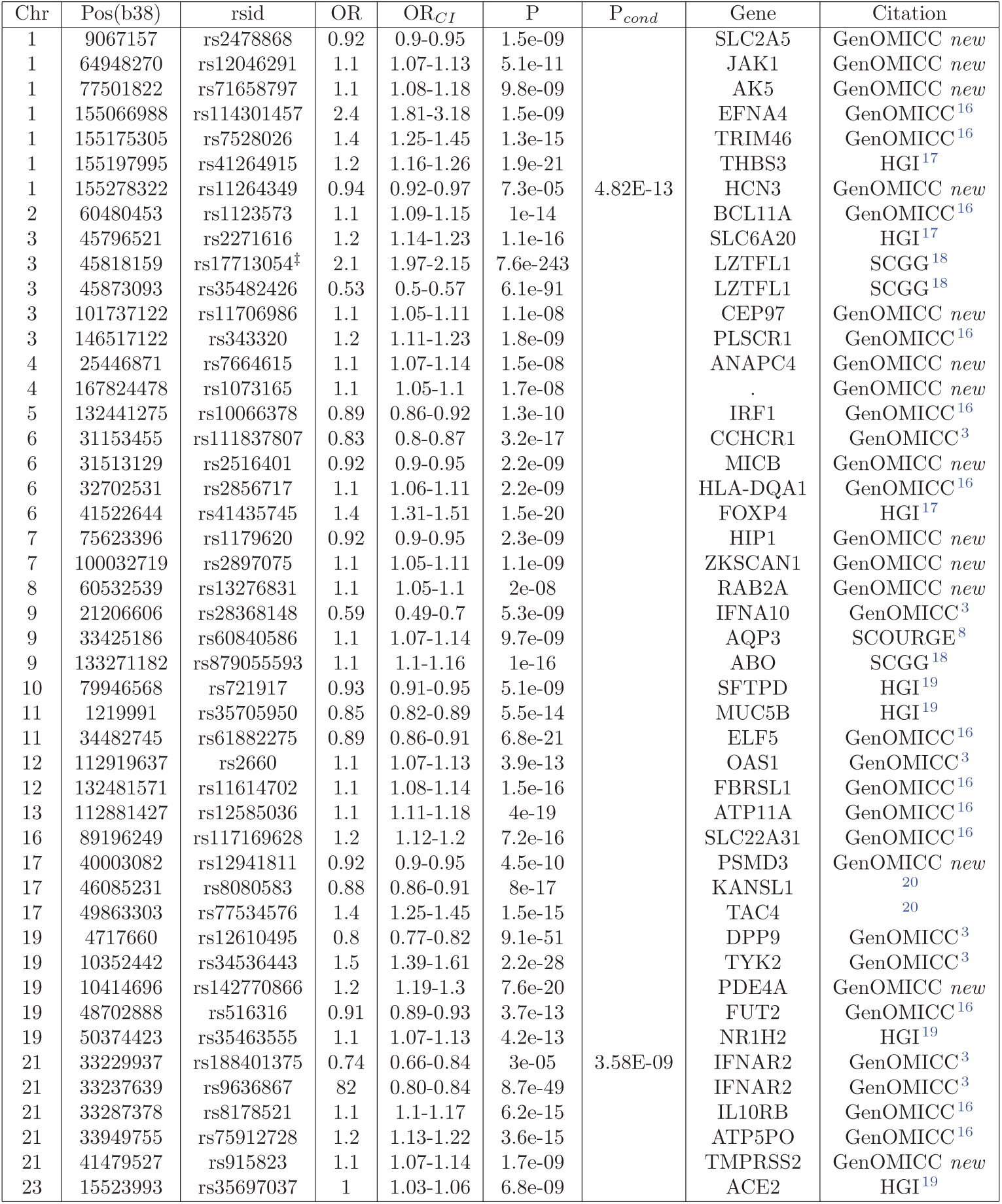
Genome-wide significant associations with critical covid. Chr: chromosome, Pos(b38): position on human genome build 38, OR: odds ratio, OR_*CI*_ : confidence interval, Gene: nearest or most plausible nearby gene, Citation: first demonstration of association. ^‡^ indicates significant heterogeneity across studies. P_*cond*_: P-value in conditional analysis in variants with P> 5 × 10^−8^ Details about conditional analysis are in Supplementary Table 3

Since no replication cohorts are available for these meta-analysis, we use heterogeneity across studies to assess the reliability of individual findings (see Supplementary Table 2 and 4). Due to the unusually extreme phenotype in the GenOMICC study, there is heterogeneity detected for the strongest associations from our previous work when compared to studies with more permissive inclusion criteria. Importantly, significant heterogeneity is not detected for the new findings we report here(Supplementary Table 2).

In order to infer the effect of genetically-determined variation in gene expression on disease susceptibility, we performed a TWAS for the critical ill phenotype using gene expression data (GTExv8^21^) for two disease-relevant tissues, lung and whole blood. We found significant associations with *P* < 1 × 10^−6^ between critical Covid-19 and predicted expression in lung (25), blood (18) and all-tissue meta-analysis (90; Supplementary file TWAS.xlsx)

## Discussion

In this latest analysis of the GenOMICC study of critical Covid-19, building on the foundations of previously published data from our group and others, we report a total of 45 genetic associations with severe Covid-19, of which 14 have not been reported previously (Table 2). After conditional analysis we find two extra secondary signals in known loci (see details in Materials and methods). In this brief report we provide a superficial description of selected findings. Our results are provided in full in Supplementary Tables 2 & 4.

We find a strong association in a key intracellular signalling kinase, JAK1, which is stimulated by numerous cytokines including Type I interferons and IL-6. Along with TYK2, which we previously reported to be associated with severe Covid-19, ^3^ JAK1 is the target for JAK inhibitors, which have recently been shown to be effective therapeutics in Covid-19. ^15^ Although the direction of effect is not clear from our genetic data for either gene, the therapeutic signal is consistent across multiple trials, providing proof-of-concept for target identification using genetics in critical illness.

We report a new lead variant within CSF2, the gene encoding granulocyte-macrophage colony stimulating factor (GM-CSF), a key cytokine in production and differentiation of myeloid cells including monocytes, macrophages and neutrophils. We previously reported that circulating levels of GM-CSF are associated with severe disease, ^22^ supporting a role for therapeutic targeting in severe Covid. In addition, we show that low expression of *PDE4A* is associated with critical Covid-19. This phosphodiesterase regulates production of multiple inflammatory cytokines by myeloid cells and is targeted by several existing drugs for treatment of a range of inflammatory diseases. ^23;24^

Multiple genes implicated in viral entry are associated with severe disease. In addition to ACE2, ^19^ we see for the first time a genome-wide significant association in TMPRSS2, a key host protease which facilitates viral entry, which we have previously studied as a candidate gene. ^25^ This effect may be lineage-specific. ^26^ A strong GWAS association is seen in RAB2A (Table 2), with TWAS evidence more expression of this gene is associated with worse disease (Table 3). This gene is highly ranked in our previous MAIC ^27^ meta-analysis of host genes implicated in SARS-CoV-2 interaction using in vitro and clinical data, ^28^ and is consistent with CRISPR screen evidence showing that RAB2A is required for viral replication. ^29^

**Table 3:** Selected TWAS results for lung, whole blood and metaTWAS with *Z* > 5 in any analysis and *P*_*meta*_ < 1×10^−5^ (See Supplementary File twas.xlsx for full results). Gene: Gene symbol, *Z*_*lung*_: TWAS Z-score in lung, *Z*_*blood*_: TWAS Z-score in blood, *Z*_*meta*_: metaTWAS mean Z-score, *Z*_*SD*_: metaTWAS Z-score standard deviation, *n*_*meta*_: number of SNPs included in metaTWAS, *P*_*meta*_: metaTWAS P-value.

| Gene | $Z_{lung}$ | $Z_{blood}$ | $Z_{meta}$ | $Z_{SD}$ | $n_{meta}$ | $P_{meta}$ |
| --- | --- | --- | --- | --- | --- | --- |
| CCR9 |  | 31 | 13 | 11 | 7 | 2.2e-224 |
| SLC6A20 | 5.7 |  | 9.2 | 6.2 | 33 | 6.9e-224 |
| IL10RB | 8.4 | -3.1 | 7.1 | 3.9 | 46 | 4e-33 |
| CCR2 | 1.6 | 0.51 | 5.9 | 7.7 | 35 | 1.3e-42 |
| TRIM46 |  |  | 5.8 | 5.1 | 18 | 4.7e-22 |
| CDC37 |  |  | 5.1 | 0.92 | 27 | 3.5e-08 |
| MUC1 | 5.7 | 7 | 5 | 2.7 | 42 | 8.2e-24 |
| RAB2A | 5 | 3.9 | 4.6 | 0.88 | 45 | 1.3e-06 |
| TCF19 | 5.6 | 4.6 | 4.4 | 2.7 | 49 | 1.1e-10 |
| FYCO1 | 8.9 | 3.7 | 4.2 | 4.1 | 47 | 2e-41 |
| GSDMA | 5.9 | 0.56 | 3.8 | 2.2 | 38 | 3.3e-06 |
| ELF5 | 5.8 |  | 3.7 | 1.7 | 9 | 2.5e-08 |
| ABO | 5.1 | -1.7 | 3.7 | 1.9 | 48 | 1.6e-07 |
| CCHCR1 | 4.3 | 4.3 | 3.5 | 2.1 | 49 | 2.9e-12 |
| ATP5O | 6.4 | 2.4 | 3.3 | 1.6 | 47 | 3.8e-11 |
| RP11-387H17.6 | 4.3 |  | 2.7 | 2.8 | 5 | 8.5e-06 |
| CAT | 4.1 | 3.1 | 2.7 | 2.2 | 49 | 5.5e-13 |
| TYK2 | 5.8 | 7.8 | 2.7 | 3.7 | 46 | 8.7e-17 |
| HLA-C | 4.6 | 5.9 | 2.7 | 2.5 | 48 | 2.2e-14 |
| ADAM15 | -6.4 | 3 | 2.5 | 2.1 | 46 | 8.9e-16 |
| GSTCD | -4.7 | 2.8 | 2.3 | 2.4 | 47 | 7.6e-06 |
| MED24 | -1.4 | 4.9 | 2.2 | 3.4 | 46 | 8.9e-08 |
| HIP1 | 4.1 | -5.8 | 0.94 | 1.7 | 42 | 1.8e-07 |
| OAS3 | 0.97 | 4.5 | 0.75 | 4.1 | 37 | 9.7e-09 |
| ZNF778 | 4.2 | 0.24 | 0.74 | 2.1 | 49 | 1.7e-06 |
| FUT2 | 6.6 |  | 0.36 | 5.2 | 28 | 2e-10 |
| FOXP4 | 8.7 |  | 0.12 | 2 | 28 | 8e-16 |
| ARHGAP27 | -6.6 | 3.6 | 0.098 | 5.2 | 44 | 7.8e-10 |
| EFNA1 | -2.5 | -6.6 | 0.042 | 2.6 | 48 | 2.2e-07 |
| ACSL6 | 3.1 | 4.6 | -0.41 | 3.6 | 44 | 1.4e-07 |
| RAVER1 | -3.8 | 5.7 | -0.56 | 2 | 45 | 7.2e-10 |
| XCR1 | -2.7 | 5.7 | -0.7 | 6.1 | 23 | 1.6e-122 |
| JAK1 | -5.3 | 0.82 | -1.7 | 3.3 | 47 | 6.9e-06 |
| CEP97 | -5.3 | 0.86 | -1.7 | 2.8 | 17 | 8.8e-06 |
| MAMSTR | -1.7 | 5.5 | -1.8 | 3.5 | 45 | 9.5e-09 |
| NXPE3 | -3.5 | -4.3 | -2.2 | 2 | 47 | 4.2e-06 |
| PSORS1C2 | -4.3 |  | -2.4 | 1.7 | 39 | 4.6e-10 |
| IFNAR1 | -4.1 | -3.6 | -2.5 | 2.2 | 49 | 1.5e-06 |
| OAS1 | -4.7 | -3.3 | -2.9 | 1.8 | 41 | 2.7e-09 |
| NTN5 | -4.3 | 3.7 | -3.4 | 3.1 | 47 | 9.8e-09 |
| CCR5 | -15 | 2.1 | -3.7 | 3.5 | 46 | 9.3e-47 |
| DPP9 | -1.2 | -5.5 | -3.9 | 3.8 | 46 | 4.3e-41 |
| CSF3 | -5.6 |  | -4 | 2 | 38 | 7e-07 |
| LINC01301 | -5 |  | -4.2 | 1.2 | 38 | 4.4e-06 |
| IFNAR2 | -5.4 | -4.1 | -4.4 | 7 | 48 | 6.7e-52 |
| RASIP1 | -5.2 | -5.2 | -4.6 | 2 | 32 | 3.7e-08 |
| WNT3 | -7.6 | -7 | -4.7 | 2.3 | 47 | 1.2e-10 |
| NAPSA | -0.56 | -6.8 | -6.1 | 1.9 | 39 | 1.2e-10 |
| CXCR6 | -21 |  | -8.1 | 9.6 | 38 | 4.4e-134 |
| CCR3 | -6.2 | -1.8 | -9 | 6 | 6 | 2e-65 |

TWAS provides a broad assessment of the effect on illness severity of genotype-predicted expression of specific transcripts in relevant tissues. TWAS results suggest a range of strongly-plausible genes, with direction of effect for gene expression effects in lung, whole blood and cross-tissue metaTWAS (Table 3, Supplemental file twas.xlsx). These include intriguing opposing effect estimates for predicted expression of various chemokine receptors (CCR1, CCR2, CCR9 vs CCR3 and CCR5) interferon-*α* subtypes (IFNA10 vs IFNA8) and intercellular adhesion molecules (ICAM1 vs ICAM3, ICAM5). Caution is needed in interpretation of these expression effects since the molecular mechanism underlying association is not known.

Although our focus on critical illness enhances discovery power, it has the disadvantage of combining genetic signals for multiple stages in disease progression including viral exposure, infection and replication, and development of inflammatory lung disease. From these data alone we cannot identify when in disease progression, or where in the body, the causal effect happens.

Because we performed a meta-analysis of multiple studies which may have slightly different definitions of the phenotype, effect sizes differ between studies. This, together with ancestry-specific effects, ^3^ may explain heterogeneity in strong GWAS signals, such as the LZTFL1 signal in table 2. Different studies also have sets of variants which are not completely overlapping, causing P-values between variants in high LD to be more different than expected. Although most of the studies contain individuals from multiple ancestries, a large majority of the individuals are from European ancestry. In future work, there is a scientific and moral imperative to include the full diversity of human populations.

Together, these results deepen our understanding of disease pathogenesis and highlights new biological mechanisms of disease, some of which have the potential for therapeutic targeting.

## Supporting information

TWAS.xlsx

## Data Availability

All data, including downloadable summary data and access applications for individual-level data, can be obtained through the GenOMICC gateway site https://genomicc.org/data.

https://genomicc.org/data

## Acknowledgements

We thank the patients and their loved ones who volunteered to contribute to this study at one of the most difficult times in their lives, and the research staff in every intensive care unit who recruited patients at personal risk during the most extreme conditions ever witnessed in most hospitals. GenOMICC was funded by Sepsis Research (the Fiona Elizabeth Agnew Trust), the Intensive Care Society, a Wellcome Trust Senior Research Fellowship (J.K.Baillie, 223164/Z/21/Z), the Department of Health and Social Care (DHSC), Illumina, LifeArc, the Medical Research Council, UKRI, a BBSRC Institute Program Support Grant to the Roslin Institute (BBS/E/D/20002172, BBS/E/D/10002070 and BBS/E/D/30002275) and UKRI grants MC PC 20004, MC PC 19025, MC PC 1905, and MRNO2995X/1. This research is supported in part by the Data and Connectivity National Core Study, led by Health Data Research UK in partnership with the Office for National Statistics and funded by UK Research and Innovation (grant ref MC PC 20029). We acknowledge NHS Digital, Public Health England and the Intensive Care National Audit and Research Centre who provided clinical data on the participants. This study owes a great deal to the National Institute for Healthcare Research Clinical Research Network (NIHR CRN) and the Chief Scientist’s Office (Scotland), who facilitate recruitment into research studies in NHS hospitals, and to the global ISARIC and InFACT consortia. GenOMICC genotype controls were obtained using UK Biobank Resource under project 788 funded by Roslin Institute Strategic Programme Grants from the BBSRC (BBS/E/D/10002070 and BBS/E/D/30002275) and Health Data Research UK (references HDR-9004 and HDR-9003). The work of LK was supported by an RCUK Innovation Fellowship from the National Productivity Investment Fund (MR/R026408/1).

We would like to thank the research participants and employees of 23andMe for making this work possible. A full list of contributors who have provided data that was collated in the HGI project, including previous iterations, is available at https://www.covid19hg.org/acknowledgements.

## Contributions

EP-C, KR, KM, SK, MJC, CPP, JFW, VV, AL, EP, RC-G, AC, AF, LMu, KRo, ALa, SCH and JKB contributed to design. EP-C, KR, LK, AK, AR, AS, CAO, SW, LM, ALa, SCH and JKB contributed to data analysis. EP-C, KR, AK, AR, JM, CDR, AS, CAO, SW, LM, ALa and SCH contributed to bioinformatics. EP-C, KR, AK, JM, CDR, RT, AN, MGS, MJC, LM, SCH and JKB contributed to writing and reviewing the manuscript. SW, FG and WO contributed to project management. FG, WO, KM, SK, AN, MGS, JK, MS-H, CS, CH, PH, LL, DM, HM, PJO, TW, AT, RHS, CPP, AF, LMu, KRo, ALa and SCH contributed to oversight. FG, WO and JKB contributed to ethics and governance. KM, AF and LMu contributed to sample handling and sequencing. MJC and JKB contributed to scientific leadership. CPP, KRo, SCH and JKB contributed to conception. CPP, JFW, VV, AL, EP, RC-G, AC and KRo contributed to reviewing the manuscript. KRo and ALa contributed to clinical data management.

## Code availability

Code to calculate the imputation of *P* -value based on LD SNPs is available at https://github.com/baillielab/GenOMICC_GWAS

## Materials and methods

### Ethical approval

All participants, or their representatives where appropriate, gave informed consent. Details of ethical review can be found in Table 4.

**Table 4:**
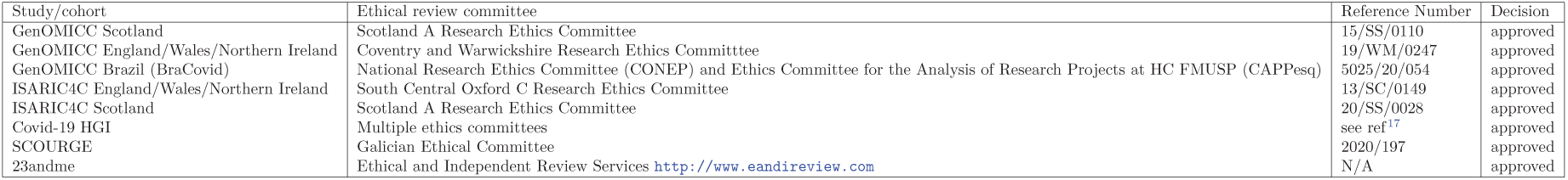
Ethical review details for primary cohorts.

| Study/cohort | Ethical review committee | Reference Number | Decision |
| --- | --- | --- | --- |
| GenOMICC Scotland | Scotland A Research Ethics Committee | 15/SS/0110 | approved |
| GenOMICC England/Wales/Northern Ireland | Coventry and Warwickshire Research Ethics Committee | 19/WM/0247 | approved |
| GenOMICC Brazil (BraCovid) | National Research Ethics Committee (CONEP) and Ethics Committee for the Analysis of Research Projects at HC FMUSP (CAPPesq) | 5025/20/054 | approved |
| ISARIC4C England/Wales/Northern Ireland | South Central Oxford C Research Ethics Committee | 13/SC/0149 | approved |
| ISARIC4C Scotland | Scotland A Research Ethics Committee | 20/SS/0028 | approved |
| Covid-19 HGI | Multiple ethics committees | see ref<sup>17</sup> | approved |
| SCOURGE | Galician Ethical Committee | 2020/197 | approved |
| 23andme | Ethical and Independent Review Services http://www.eandireview.com | N/A | approved |

### Hospitalisation meta-analysis

The hospitalised phenotype includes patients who were hospitalised with a laboratory-confirmed SARS-Cov2 infection. In this analysis we included GenOMICC, GenOMICC Brazil, ISARIC4C, HGIv6 B2 phenotype with subtraction of GenOMICC data, SCOURGE hospitalised vs population and mild cases, and 23andme broad respiratory phenotype. Summary description of each analysis can be found in Supplementary material and a table with the included studies can be found in Supplementary Table 1.

### Critical illness meta-analysis

The critically ill Covid-19 group included patients who were hospitalized owing to symptoms associated with laboratory-confirmed SARS-CoV-2 infection and who required respiratory support or whose cause of death was associated with Covid-19 In the critical analysis we included GenOMICC, critically-ill patients from ISARIC4C, HGIv6 phenothype A2 with subraction of GenOMICC data, SCOURGE severity grades 3 and 4 vs population controls, and 23andme respiratory support phenotype. Summary description of each analysis can be found in Supplementary material and a table with the included studies can be found in table 1.

### Meta-analysis

All meta-analysis across studies were performed using an inverse-variance weighting method and control for population stratification in the METAL software ^30^. Variants were filtered out if they were not present in one of the three biggest studies: GenOMICC European ancestry, HGIv6, or SCOURGE. Allele frequency was calculated as the average frequency across studies with the METAL option AVERAGEFREQ. Heterogeneity was calculated using a Cochran’s Q-test implemented in METAL. For variants in the same position with different REF and ALT alleles across studies, the GenoMICC variant in European population was selected and the rest were removed. Finally, variants with switched ALT and REF alleles between HGIv6 and GenOMICC were also removed based on differences in allele frequency of the alternative allele. Variants were annotated to the closest genes using dbsnp version b151 GRCh38p7 and biomaRt R package version 2.46.3. ^31^.

### LD clumping

We used plink 1.9 to clump variants that were genome-wide significant for each analysis with *P* 1 = 5 × 10^−8^, *P* 2 = 0.01, clump distance 1500Kb and *r*^2^ = 0.1. As a reference population for clumping we used individuals from European ancestry with whole genome sequence available in the GenOMICC study and whole genomes from 100K Genomics England project ^4^.

### Conditional analysis

We performed a step-wise conditional analysis to find independent signals. In the hospitalised analysis we performed an European-specific meta-analysis to find conditionally independent signals. For the critical illness meta-analysis European-specific data is not available, but European ancestry is largely predominant (87.2% of critically-ill cases) and we performed the conditional analysis using the meta-analysis results of the whole cohort. To perform the conditional analysis we used GCTA 1.9.3 –cojo-slct function ^32^. The parameters for the function were *pval* = 5 × 10^−8^, a distance of 10,000 kb and a colinear threshold of 0.9^33^, and the reference population for the conditional analysis were individuals from European ancestry with whole genome sequence available in the GenOMICC study and whole genomes from 100K Genomics England project ^4^.

### Transcriptome-wide Association Studies

We performed TWAS in the MetaXcan framework and the GTExv8 eQTL and sQTL MASHR-M models available for download in http://predictdb.org/. We first calculated individual TWAS for whole blood and lung with the S-PrediXcan function ^34;35^. Then we performed a metaTWAS including data from all tissues to increase statistical power using s-MultiXcan ^36^. We applied Bonferroni correction to the results in order to choose significant genes and introns for each analysis.

## References

[1] Horby, P. et al. Dexamethasone in Hospitalized Patients with Covid-19 — Preliminary Report. New England Journal of Medicine (2020).

[2] Dorward, D. A. et al. Tissue-specific Immunopathology in Fatal COVID-19. American Journal of Respiratory and Critical Care Medicine (2020).

[3] Pairo-Castineira, E. et al. Genetic mechanisms of critical illness in covid-19. Nature 591, 92–98 (2021).

[4] Kousathanas, A. et al. Whole genome sequencing reveals host factors underlying critical Covid-19. Nature 1–10 (2022).

[5] Abani, O. et al. Tocilizumab in patients admitted to hospital with COVID-19 (RECOVERY): A randomised, controlled, open-label, platform trial. The Lancet 397, 1637–1645 (2021).

[6] Baillie, J. K. Targeting the host immune response to fight infection. Science 344, 807–808 (2014).

[7] Docherty, A. B. et al. Features of 20 133 UK patients in hospital with covid-19 using the ISARIC WHO Clinical Characterisation Protocol: Prospective observational cohort study. BMJ 369 (2020).

[8] Carracedo, Á. & on Covid-19 (scourge), S. C. t. U. R. o. h. G. A genome-wide association study of COVID-19 related hospitalization in Spain reveals genetic disparities among sexes (2021).

[9] Docherty, A. B. et al. Features of 20 133 uk patients in hospital with covid-19 using the isaric who clinical characterisation protocol: prospective observational cohort study. BMJ (Clinical research ed.) 369, m1985 (2020).

[10] Niemi, M. E. K. et al. Mapping the human genetic architecture of covid-19. Nature 600, 472–477 (2021).

[11] Horby, P., Nguyen, N. Y., Dunstan, S. J. & Baillie, J. K. The Role of Host Genetics in Susceptibility to Influenza: A Systematic Review. PLoS ONE 7, e33180 (2012).

[12] Patarčić, I. et al. The role of host genetic factors in respiratory tract infectious diseases: Systematic review, meta-analyses and field synopsis. Scientific Reports 5 (2015).

[13] Zhou, S. et al. A neanderthal oas1 isoform protects individuals of european ancestry against covid-19 susceptibility and severity. Nature medicine 27, 659–667 (2021).

[14] Wickenhagen, A. et al. A prenylated dsrna sensor protects against severe covid-19. Science (New York, N.Y.) 374, eabj3624 (2021).

[15] Horby, P. W. et al. Baricitinib in patients admitted to hospital with COVID-19 (RECOVERY): A randomised, controlled, open-label, platform trial and updated meta-analysis (2022).

[16] Kousathanas, A. et al. Whole genome sequencing identifies multiple loci for critical illness caused by covid-19. medRxiv (2021). URL https://www.medrxiv.org/content/early/2021/09/08/2021.09.02.21262965.

[17] COVID-19 Host Genetics Initiative. Mapping the human genetic architecture of COVID-19. Nature (2021). URL https://doi.org/10.1038/s41586-021-03767-x.

[18] Ellinghaus, D. et al. Genomewide association study of severe covid-19 with respiratory failure. The New England journal of medicine 383, 1522–1534 (2020).

[19] Initiative, C.-. H. G. & Ganna, A. Mapping the human genetic architecture of COVID-19: An update (2021).

[20] Degenhardt, F. et al. New susceptibility loci for severe COVID-19 by detailed GWAS analysis in European populations (2021).

[21] Consortium, T. G. The GTEx Consortium atlas of genetic regulatory effects across human tissues. Science 369, 1318–1330 (2020). URL https://science.sciencemag.org/content/369/6509/1318. Publisher: American Association for the Advancement of Science eprint:https://science.sciencemag.org/content/369/6509/1318.full.pdf.

[22] Thwaites, R. S. et al. Inflammatory profiles across the spectrum of disease reveal a distinct role for GM-CSF in severe COVID-19. Science Immunology 6 (2021).

[23] Sakkas, L. I., Mavropoulos, A. & Bogdanos, D. P. Phosphodiesterase 4 inhibitors in immune-mediated diseases: Mode of action, clinical applications, current and future perspectives. Current medicinal chemistry 24, 3054–3067 (2017).

[24] Contreras, S., Milara, J., Morcillo, E. & Cortijo, J. Selective inhibition of phosphodiesterases 4A, b, c and d isoforms in chronic respiratory diseases: Current and future evidences. Current pharmaceutical design 23, 2073–2083 (2017).

[25] David, A. et al. A common TMPRSS2 variant has a protective effect against severe COVID-19. Current Research in Translational Medicine 70, 103333 (2022).

[26] Meng, B. et al. Altered TMPRSS2 usage by SARS-CoV-2 Omicron impacts tropism and fusogenicity. Nature 1–1 (2022).

[27] Li, B. et al. Genome-wide CRISPR screen identifies host dependency factors for influenza A virus infection. Nature Communications 11, 164 (2020).

[28] Parkinson, N. et al. Dynamic data-driven meta-analysis for prioritisation of host genes implicated in COVID-19. Scientific Reports 10, 22303 (2020).

[29] Hoffmann, H.-H. et al. Functional interrogation of a sars-cov-2 host protein interactome identifies unique and shared coronavirus host factors. bioRxiv : the preprint server for biology (2020).

[30] Willer, C. J., Li, Y. & Abecasis, G. R. METAL: fast and efficient meta-analysis of genomewide association scans. Bioinformatics (Oxford, England) 26, 2190–2191 (2010).

[31] Durinck, S., Spellman, P. T., Birney, E. & Huber, W. Mapping identifiers for the integration of genomic datasets with the r/bioconductor package biomart. Nature protocols 4, 1184–91 (2009).

[32] Yang, J., Lee, S. H., Goddard, M. E. & Visscher, P. M. Gcta: a tool for genome-wide complex trait analysis. American journal of human genetics 88, 76–82 (2011). URL https://pubmed.ncbi.nlm.nih.gov/21167468/.

[33] Yang, J. et al. Conditional and joint multiple-SNP analysis of GWAS summary statistics identifies additional variants influencing complex traits. Nature Genetics 44, 369–375 (2012). URL https://doi.org/10.1038/ng.2213.

[34] Barbeira, A. N. et al. Exploring the phenotypic consequences of tissue specific gene expression variation inferred from GWAS summary statistics. Nature Communications 9, 1825 (2018).

[35] Gamazon, E. R. et al. A gene-based association method for mapping traits using reference transcriptome data. Nature Genetics 47, 1091–1098 (2015).

[36] Barbeira, A. N. et al. Integrating predicted transcriptome from multiple tissues improves association detection. PLOS Genetics 15, 1–20 (2019).

