## Supplementary Information for "GWAS and meta-analysis identifies multiple new genetic mechanisms underlying severe Covid-19"

**References** 22

### Contributing Studies

#### GenOMICC and ISARIC4C

Patients were recruited to the GenOMICC (Genetics Of Mortality In Critical Care) study in 224 UK intensive care units (https://genomicc.org). All cases had confirmed Covid-19 according to local clinical testing and were deemed, in the view of the treating clinician, to require continuous cardiorespiratory monitoring. In UK practice this kind of monitoring is undertaken in high-dependency or intensive care units. Additional Covid-19 confirmed hospitalised cases were recruited through the International Severe Acute Respiratory Infection Consortium (ISARIC) Coronavirus Clinical Characterisation Consortium (4C). Current and previous versions of the study protocol are available at https://genomicc.org/protocol/. All participants gave informed consent.
Participants were recruited to the mild Covid-19 cohort on the basis of having experienced mild (non-hospitalised) or asymptomatic Covid-19. Participants volunteered to take part in the study via a microsite and were required to self-report the details of a positive Covid-19 test. Volunteers were prioritised for genome sequencing based on demographic matching with the critical Covid-19 cohort considering self-reported ancestry, sex, age and location within the UK

##### Whole genome sequencing summary statistics

8794 critically ill individuals and 1809 mild Covid-19 controls were sequenced through the GenOMICC study. To increase control numbers, we added general population controls from 100k-genomes cohort. Participants were enrolled in the 100,000 Genomes Project from families with a broad range of rare diseases, cancers and infection by 13 regional NHS Genomic Medicine Centres across England and in Northern Ireland, Scotland and Wales. For this analysis, participants for whom a positive SARS-CoV-2 test had been recorded as of March, 2021 were not included due to uncertainty in the severity of Covid-19 symptoms. Only participants for whom genome sequencing was performed from blood derived DNA were included and participants with haematological malignancies were excluded to avoid potential tumour contamination. DNA extraction, sequencing, ancestry estimation, kinship estimation, principal components calculation and QC pipelines were performed as described in Kousathanas et al.[1]
GWAS was performed using a 2-step logistic mixed model regression approach as implemented in SAIGE v0.44.5 for single variant association analyses with sex, age, age squared , age by sex interaction and 20 principal components as covariates. The principal components were computed separately by predicted genetic ancestry (i.e, EUR-specific, AFR-specific, etc.), to capture subtle structure effects.

##### Genotype summary statistics

8143 Critically ill cases recruited through the GenOMICC, and 888 additional Covid-19 confirmed hospitalised cases recruited through ISARIC4C were genotyped. DNA extraction, sample QC, genotype QC, kinship estimation, ancestry estimation and imputation were performed with the pipelines described in Pairo-Castineira et al.[2].
After these steps and removing individuals with whole genome sequencing data available[1] there were 1900 unrelated individuals assigned to the European ancestry group, 259 unrelated individuals assigned to the South Asian ancestry group, and 144 unrelated individuals assigned to the African ancestry group. Individuals from East Asian and American ancestries were removed from the analysis because there were less than 100 cases for each of them.
Principal components were calculated as in our previous work[2] for all GenOMICC participants and UK Biobank individuals, as well as, for each ancestry group individually and are illustrated in Supplementary Figure 1 and Supplementary Figure 2 respectively.
UK Biobank participants from a subset with no relationships up to third degree were considered as potential controls if they were not identified by the UK Biobank as outliers based on either genotyping missingness rate or heterogeneity, and their sex inferred from the genotypes matched their self-reported sex. After excluding participants who had received PCR tests for Covid-19, based on information downloaded from the UK Biobank on 27/01/2022. For each GenOMICC individual we considered the subset of UK Biobank individuals with matching assigned ancestry group and sex, and selected the five closest, based on euclidean distance in the space of the first six ancestry specific genomic principal components, as controls.

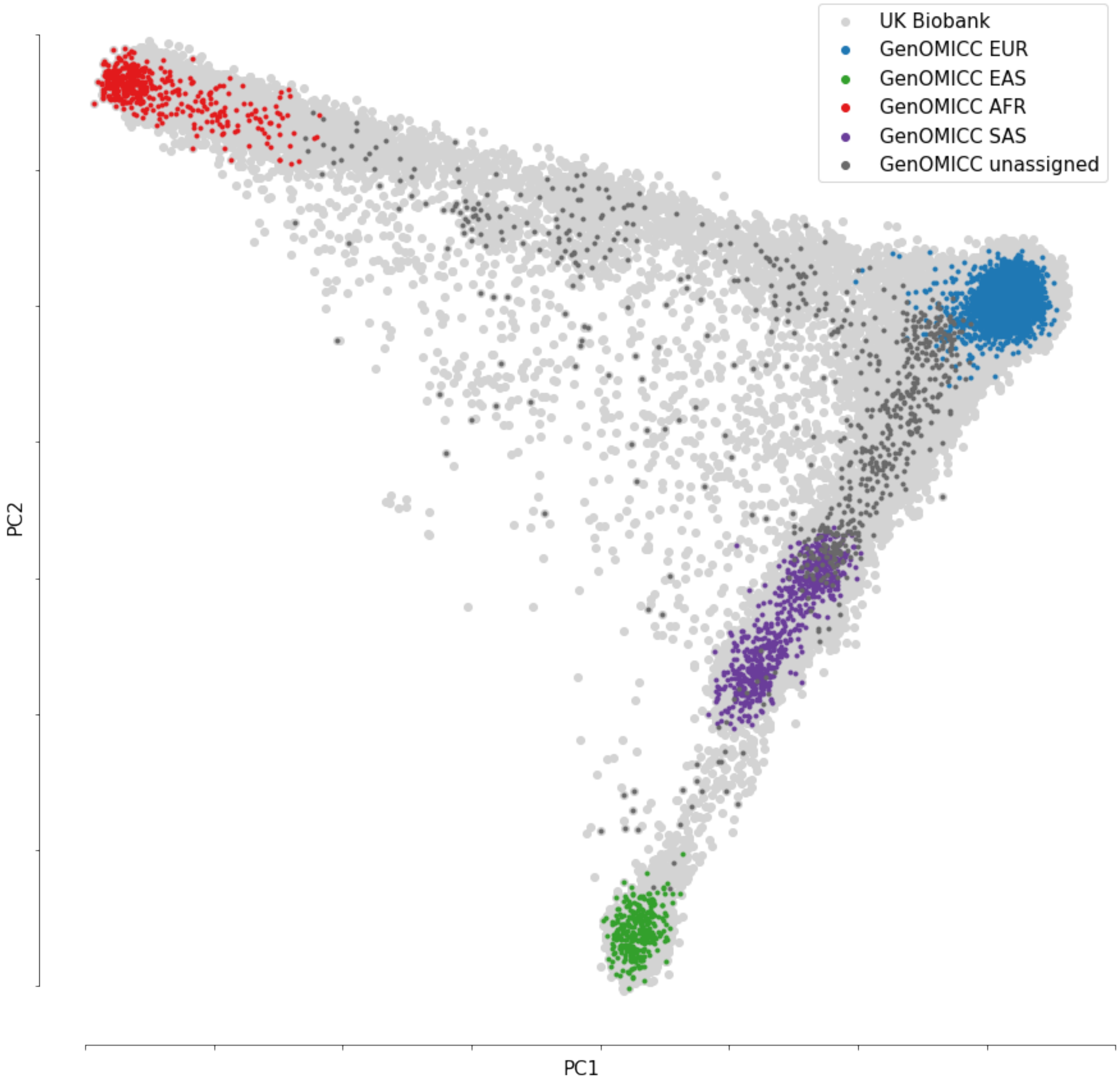

Figure 1: Genomic PCA plot showing the first two principal components for the combination of all UK Biobank and GenOMICC participants. Assigned ancestry group for GenOMICC participants is indicated through colour.

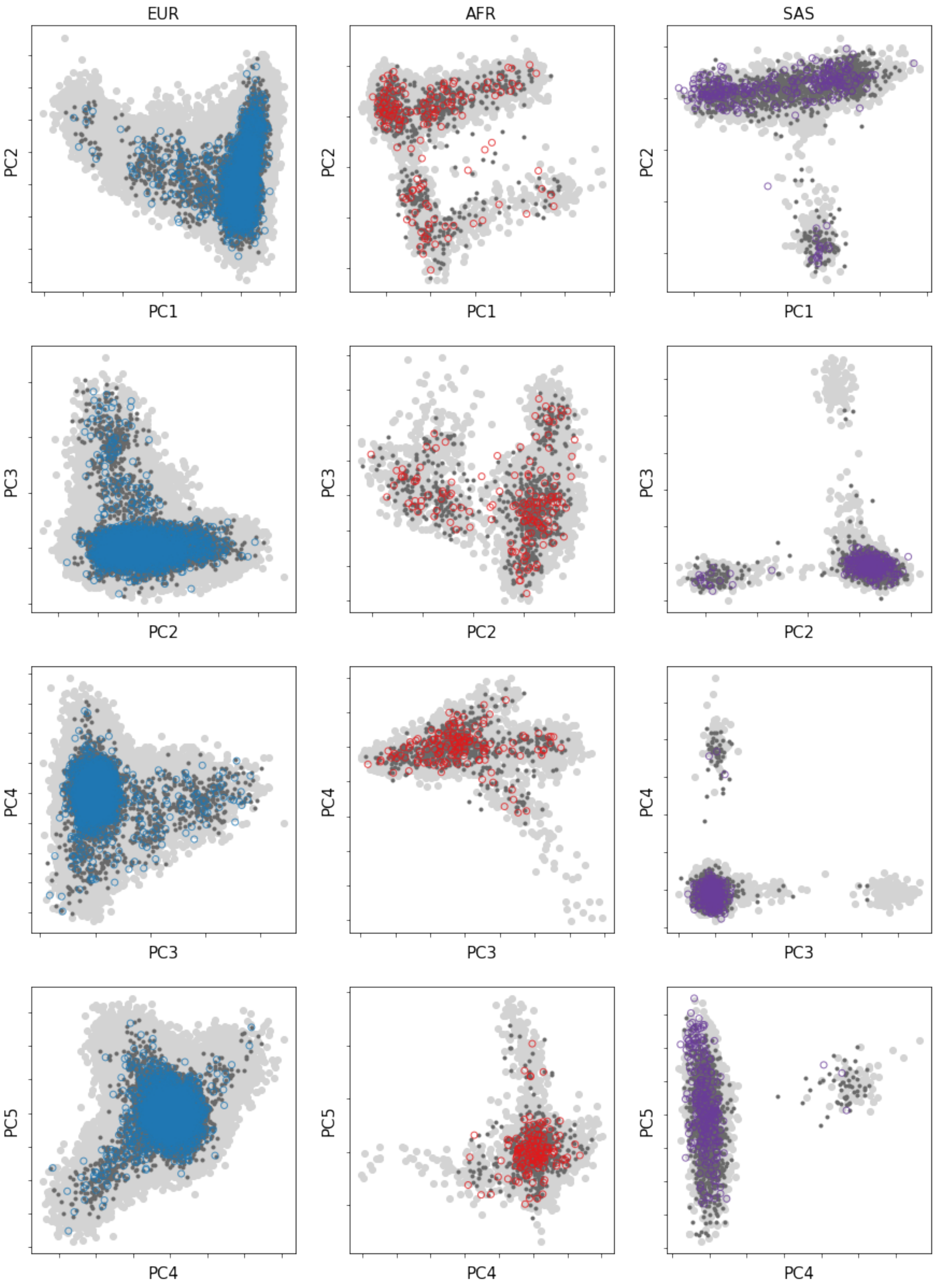

Figure 2: Genomic PCA plots showing the distribution of all cases and controls for the first 5 principal components for each ancestry group. Cases are shown as coloured open circles: European (EUR, blue), African (AFR, red), and South Asian (SAS, purple). Controls are dark grey closed circles. UK Biobank population background is shown as light grey closed circles.

Test for association between case–control status and allele dosage at the was performed by fitting a logistic regression model using PLINK 2.00 with sex, age, mean-centred age2, deprivation score decile of residential postcode, and the first 10 ancestry specific genomic principal components as covariates. GWAS results for individuals from European ancestry were filtered for $MAF > 0.01$, $HWEP-value > 10^{-50}$, and genotyping rate $> 0.99$ and imputation score $> 0.9$ in both GenOMICC and UK Biobank. To avoid bias for using a different genotyping array and imputation method between cases and controls MAF for each SNP was compared between UK Biobank and gNOMAD non-Finnish European individuals, and SNPs were removed from following two rules (1)In SNPs with MAF $> 0,1$ in gnomAD, an absolute difference in MAF of 0.05 between gnomAD and UK Biobank controls; (2) in SNPs with MAF $< 0.1$ in gnomAD, a difference in MAF of $> 0.25 \times MAF_{gnomad}$ between UK Biobank controls and gnomAD. After this filter, a further LD-check filter, following Kousathanas et al[1] was used to clean the GWAS from false positives. GWAS analyses of individuals of South Asian and African ancestries were filtered for variants filtered for a MAF in UK Biobank corresponding to the same ancestry $> 0.05$ and then for the SNPs that passed quality control in the European GWAS. For the Critical-illness GWAS, ISARIC4C hospitalised individuals that did not fulfill the critical illness criteria were removed from the analysis with their matched controls, leaving 101 Covid-19 critically-ill cases from African ancestry, 200 critically-ill cases from South Asian Ancestry and 1487 critically-ill cases from European ancestry and 5 matched controls for each case.

##### BraCovid/GenoMICC Brazil

Covid-19 patients were enrolled after hospitalisation in one of the following tertiary care centers in the metropolitan area of Sao Paulo, Brazil: Instituto do Coração, and Instituto Central do Hospital das Clínicas da Faculdade de Medicina da Universidade de São Paulo. Non-hospitalised cases were selected by serological studies surveys for previous SARS-CoV-2 infection or SARS-CoV-2 PCR test among health professionals or the general population.After signing an informed consent, a sample of whole-blood already collected for in-hospital biochemical analysis or SARS-CoV-2 serology was used for genomic DNA extraction.DNA extraction, genotyping,imputation and QC pipelines are described in Pereira et al.[3]
GWAS of Covid-19 hospitalised cases versus Covid-19 positive mild controls was performed using a logistic regression model in PLINK 2.0 including age, sex, age, array type and four principal components as covariates. Due to the level of admixture and complex genetic structure present in the Brazilian population, three different ancestry subgroups were defined (European, African, Native American), and a fixed-effect meta-analysis was calculated using the plink meta-analysis routine[3].

#### SCOURGE

In Spain, 11,939 Covid-19 positive cases were recruited as part of SCOURGE study from 34 centres in 25 cities.[4] Study samples and data were collected by the participating centers, through their respective biobanks after informed consent, with the approval of the respective Ethic and Scientific Committees. The whole project was approved by the Galician Ethical Committee Ref 2020/197. Individuals were diagnosed as Covid-19 positive through a PCR-based test or according to local clinical and laboratory procedures. All cases were classified in a five-level severity scale.Two Spanish sample collections with unknown Covid-19 status were included as general population controls in some analyses: 3,437 samples from the Spanish DNA biobank (https://www.bancoadn.org) and 2,506 samples from the GR@CE consortium. DNA extraction, genotyping, imputation, ancestry estimation, kinship estimation and QC pipelines are described in Carracedo et al.[4].
GWAS was performed by fitting a logisitic regression models implemented in SAIGE. For the hospitalised meta-analysis the summary statistics of the hospitalisation vs non-hospitalised (mild cases and population controls) analysis were used. For the critical illness meta-analysis, the summary statistics of severity grade 3 and 4 as defined by SCOURGE[4] vs population controls were used.

#### HGI release 6

In total 25 studies contributed to the A2 analysis, with 8779 cases and 1001572 controls and 43 studies contributed to the B2 analysis with 24274 cases and 2061529 controls without including 23andme data. All studies followed protocols approved by local Institutional Review Boards; All protocols followed local ethics recommendations and informed consent was obtained when required Meta-analysis of all cohorts was performed with and inverse-weighting variance method as described by the Covid-19 Host Genetics Initiative[5]

In order to account for signal due to sample overlap we performed a mathematical subtraction from HGIv6B2 of the GenOMICC GWAS of European genetic ancestry and the BraCovid analysis, and a mathematical subtraction from HGIv6A2 of the GenOMICC GWAS of European genetic ancestry . Publicly-available HGI data was downloaded from https://www.covid19hg.org/results/r6/. The subtraction was performed using MetaSubtract package (version 1.60) for R (version 4.0.2) after removing variants with the same genomic position and using the lambda.cohorts with genomic inflation calculated on the GenOMICC summary statistics.

#### 23andme

Participants were recruited from the customer base of 23andMe, Inc., a personal genetics company. All individuals included in the analyses provided informed consent and answered surveys online according to 23andme human subjects research protocol, which was reviewed and approved by Ethical and Independent Review Services, a private institutional review board (http://www.eandireview.com). Study participation consisted solely of web-based surveys, and different phenotypes were defined for disease severity: pneumonia, hospitalisation and respiratory support. DNA extraction, genotyping, imputation, ancestry estimation, kinship estimation and QC pipelines are described in Shelton et al.[6]. GWAS were performed for each phenotype and ancestry separately using a logistic regression using age, sex, age squared, a age:sex interaction and top ten principal components as covariates, as described in Shelton et al.[6]

For the hospitalisation meta-analysis we used summary statistics from the respiratory support or pneumonia phenotype against population controls GWAS from European, African and Latino ancestries. For the critically-ill meta-analysis we used summary statistics from the respiratory support phenotype against population controls GWAS. We filtered variants that passed internal 23andMe ancestry QC[6] and had imputation score $> 0.6$ and with MAF $> 0.005$.

### Hospitalisation meta-analysis

| Study | Dataset | Ancestry | Case definition | Control definition | Ncases | Ncontrols |
|---|---|---|---|---|---|---|
| GenOMICC | Whole-Genome Sequence | EUR | ICU admission | General population and mild cases | 5989 | 42891 |
| GenOMICC | Whole-Genome Sequence | EAS | ICU admission | General population and mild cases | 274 | 366 |
| GenOMICC | Whole-Genome Sequence | SAS | ICU admission | General population and mild cases | 788 | 3793 |
| GenOMICC | Whole-Genome Sequence | AFR | ICU admission | General population and mild cases | 440 | 1350 |
| GenomICC | Genotype | EUR | ICU admission | General population | 1347 | 6735 |
| GenomICC | Genotype | SAS | ICU admission | General population | 200 | 100 |
| GenomICC | Genotype | AFR | ICU admission | General population | 101 | 505 |
| ISARIC4C | Genotype | EUR | Hospitalised | General population | 553 | 2675 |
| ISARIC4C | Genotype | SAS | Hospitalised | General population | 59 | 295 |
| ISARIC4C | Genotype | AFR | Hospitalised | General population | 43 | 215 |
| Brazil GenOMICC | Genotype | ALL | Hospitalised | Mild cases | 3533 | 1700 |
| HGiv6 | Genotype | ALL | Hospitalised | General population | 22637 | 2052436 |
| SCOURGE | Genotype | EUR | Hospitalised | General population and mild cases | 5934 | 8810 |
| 23 and me | Genotype | EUR | Respiratory support or pneumonia | General population | 1128 | 679531 |
| 23 and me | Genotype | AMR | Respiratory support or pneumonia | General population | 218 | 94318 |
| 23 and me | Genotype | AFR | Respiratory support or pneumonia | General population | 64 | 22382 |

Table 1: Studies included in the hospitalised meta-analysis with information about ancestry, case and control defintion, and number of cases and controls included in the analysis.

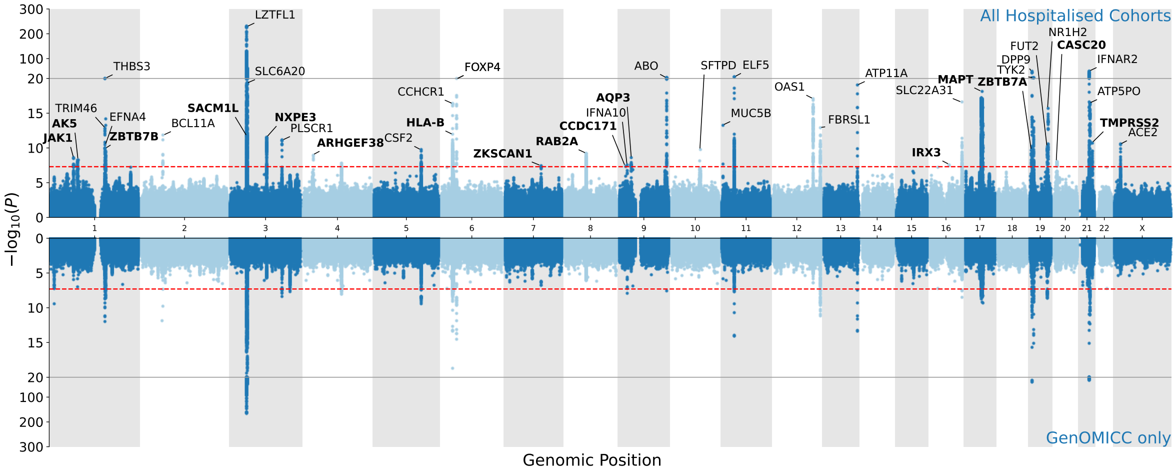

Figure 3: A Miami plot showing meta-analysis results obtained using all hospitalised phenotype cohorts (top) and using GenOMICC data only (bottom). Independent lead variants in the all hospitalised cohorts analysis are annotated with associated genes, with not previously reported associations indicated in bold.

.

| Chr | Pos(b38) | rsid | OR | $OR_{CI}$ | P | $P_{cond}$ | Het P.value | N | Gene | Citation |
|---|---|---|---|---|---|---|---|---|---|---|
| 1 | 9067157 | rs2478868 | 0.92 | 0.9-0.95 | 1.5e-09 | | 0.042 | 1754160 | SLC2A5 | GenOMICC *new* |
| 1 | 64948270 | rs12046291 | 1.1 | 1.07-1.13 | 5.1e-11 | | 0.68 | 1731600 | JAK1 | GenOMICC *new* |
| 1 | 77501822 | rs71658797 | 1.1 | 1.08-1.18 | 9.8e-09 | | 0.21 | 1748720 | AK5 | GenOMICC *new* |
| 1 | 155066988 | rs114301457 | 2.4 | 1.81-3.18 | 1.5e-09 | | 1 | 48877 | EFNA4 | GenOMICC[7] |
| 1 | 155175305 | rs7528026 | 1.4 | 1.25-1.45 | 1.3e-15 | | 0.52 | 1746390 | TRIM46 | GenOMICC[7] |
| 1 | 155197995 | rs41264915 | 1.2 | 1.16-1.26 | 1.9e-21 | | 0.67 | 1753360 | THBS3 | HGI[5] |
| 1 | 155278322 | rs11264349 | 0.94 | 0.92-0.97 | 7.3e-05 | 4.82E-13 | 0.36 | 1743390 | HCN3 | GenOMICC *new* |
| 2 | 60480453 | rs1123573 | 1.1 | 1.09-1.15 | 1e-14 | | 0.24 | 1725960 | BCL11A | GenOMICC[7] |
| 3 | 45796521 | rs2271616 | 1.2 | 1.14-1.23 | 1.1e-16 | | 0.00013 | 1016720 | SLC6A20 | HGI[5] |
| 3 | 45818159 | rs17713054‡ | 2.1 | 1.97-2.15 | 7.6e-243 | | 1.4e-12 | 1753220 | LZTFL1 | SCGG[8] |
| 3 | 45873093 | rs35482426 | 0.53 | 0.5-0.57 | 6.1e-91 | | 0.2 | 55882 | LZTFL1 | SCGG[8] |
| 3 | 101737122 | rs11706986 | 1.1 | 1.05-1.11 | 1.1e-08 | | 0.57 | 1754690 | CEP97 | GenOMICC *new* |
| 3 | 146517122 | rs343320 | 1.2 | 1.11-1.23 | 1.8e-09 | | 0.24 | 1742080 | PLSCR1 | GenOMICC[7] |
| 4 | 25446871 | rs7664615 | 1.1 | 1.07-1.14 | 1.5e-08 | | 0.28 | 1725240 | ANAPC4 | GenOMICC *new* |
| 4 | 167824478 | rs1073165 | 1.1 | 1.05-1.1 | 1.7e-08 | | 0.99 | 1754220 | . | GenOMICC *new* |
| 5 | 132441275 | rs10066378 | 0.89 | 0.86-0.92 | 1.3e-10 | | 0.036 | 1754720 | IRF1 | GenOMICC[7] |
| 6 | 31153455 | rs111837807 | 0.83 | 0.8-0.87 | 3.2e-17 | | 1.1e-06 | 1754260 | CCHCR1 | GenOMICC[2] |
| 6 | 31513129 | rs2516401 | 0.92 | 0.9-0.95 | 2.2e-09 | | 0.74 | 1752350 | MICB | GenOMICC *new* |
| 6 | 32702531 | rs2856717 | 1.1 | 1.06-1.11 | 2.2e-09 | | 0.032 | 1743290 | HLA-DQA1 | GenOMICC[7] |
| 6 | 41522644 | rs41435745 | 1.4 | 1.31-1.51 | 1.5e-20 | | 0.45 | 1725810 | FOXP4 | HGI[5] |
| 7 | 75623396 | rs1179620 | 0.92 | 0.9-0.95 | 2.3e-09 | | 0.39 | 1725980 | HIP1 | GenOMICC *new* |
| 7 | 100032719 | rs2897075 | 1.1 | 1.05-1.11 | 1.1e-09 | | 0.92 | 1753670 | ZKSCAN1 | GenOMICC *new* |
| 8 | 60532539 | rs13276831 | 1.1 | 1.05-1.1 | 2e-08 | | 0.54 | 1753710 | RAB2A | GenOMICC *new* |
| 9 | 21206606 | rs28368148 | 0.59 | 0.49-0.7 | 5.3e-09 | | 0.31 | 729815 | IFNA10 | GenOMICC[2] |
| 9 | 33425186 | rs60840586 | 1.1 | 1.07-1.14 | 9.7e-09 | | 0.035 | 936386 | AQP3 | SCOURGE[4] |
| 9 | 133271182 | rs879055593 | 1.1 | 1.1-1.16 | 1e-16 | | 0.96 | 1732540 | ABO | SCGG[8] |
| 10 | 79946568 | rs721917 | 0.93 | 0.91-0.95 | 5.1e-09 | | 0.21 | 1754240 | SFTPD | HGI[9] |
| 11 | 1219991 | rs35705950 | 0.85 | 0.82-0.89 | 5.5e-14 | | 0.68 | 1740360 | MUC5B | HGI[9] |
| 11 | 34482745 | rs61882275 | 0.89 | 0.86-0.91 | 6.8e-21 | | 0.56 | 1753250 | ELF5 | GenOMICC[7] |
| 12 | 112919637 | rs2660 | 1.1 | 1.07-1.13 | 3.9e-13 | | 0.79 | 1753650 | OAS1 | GenOMICC[2] |
| 12 | 132481571 | rs11614702 | 1.1 | 1.08-1.14 | 1.5e-16 | | 0.81 | 1754180 | FBRSL1 | GenOMICC[7] |
| 13 | 112881427 | rs12585036 | 1.1 | 1.11-1.18 | 4e-19 | | 0.26 | 1754020 | ATP11A | GenOMICC[7] |
| 16 | 89196249 | rs117169628 | 1.2 | 1.12-1.2 | 7.2e-16 | | 0.28 | 1752690 | SLC22A31 | GenOMICC[7] |
| 17 | 40003082 | rs12941811 | 0.92 | 0.9-0.95 | 4.5e-10 | | 0.45 | 1753680 | PSMD3 | GenOMICC *new* |
| 17 | 46085231 | rs8080583 | 0.88 | 0.86-0.91 | 8e-17 | | 0.73 | 1750860 | KANSL1 | [10] |
| 17 | 49863303 | rs77534576 | 1.4 | 1.25-1.45 | 1.5e-15 | | 0.33 | 1747030 | TAC4 | [10] |
| 19 | 4717660 | rs12610495 | 0.8 | 0.77-0.82 | 9.1e-51 | | 0.0019 | 1691560 | DPP9 | GenOMICC[2] |
| 19 | 10352442 | rs34536443 | 1.5 | 1.39-1.61 | 2.2e-28 | | 0.97 | 1683680 | TYK2 | GenOMICC[2] |
| 19 | 10414696 | rs142770866 | 1.2 | 1.19-1.3 | 7.6e-20 | | 0.56 | 1749100 | PDE4A | GenOMICC *new* |
| 19 | 48702888 | rs516316 | 0.91 | 0.89-0.93 | 3.7e-13 | | 0.6 | 1750800 | FUT2 | GenOMICC[7] |
| 19 | 50374423 | rs35463555 | 1.1 | 1.07-1.13 | 4.2e-13 | | 0.63 | 1073760 | NR1H2 | HGI[9] |
| 21 | 33229937 | rs188401375 | 0.74 | 0.66-0.84 | 3e-05 | 3.58E-09 | 0.4 | 734303 | IFNAR2 | GenOMICC[2] |
| 21 | 33237639 | rs9636867 | 82 | 0.80-0.84 | 8.7e-49 | | 0.12 | 1754230 | IFNAR2 | GenOMICC[2] |
| 21 | 33287378 | rs8178521 | 1.1 | 1.1-1.17 | 6.2e-15 | | 0.31 | 1726400 | IL10RB | GenOMICC[7] |
| 21 | 33949755 | rs75912728 | 1.2 | 1.13-1.22 | 3.6e-15 | | 0.1 | 1754110 | ATP5PO | GenOMICC[7] |
| 21 | 41479527 | rs915823 | 1.1 | 1.07-1.14 | 1.7e-09 | | 0.99 | 1754710 | TMPRSS2 | GenOMICC *new* |
| 23 | 15523993 | rs35697037 | 1 | 1.03-1.06 | 6.8e-09 | | 0.38 | 1723360 | ACE2 | HGI[9] |

Table 2: Full results for critical covid GWAS. Chr: chromosome, Pos(b38): position on human genome build 38, Allele1: effect allele, Allele2: other allele, OR: odds ratio, $OR_{CI}$: confidence interval, N: number of individuals in the analysis Het P.value: Heterogeneity P-value Gene: nearest or most plausible nearby gene, Citation: first demonstration of association. ‡ indicates significant heterogeneity across studies. $P_{cond}$: P-value in conditional analysis in variants with $P > 5 \times 10^{-8}$

| Chr | Pos(b38) | rsid | Allele1 | Allele2 | beta | SE | P | $beta_{cond}$ | $SE_{cond}$ | $P_{cond}$ | $LD_r$ | Gene |
|---|---|---|---|---|---|---|---|---|---|---|---|---|
| 1 | 9067157 | rs2478868 | A | C | -0.0785 | 0.013 | 1.46E-09 | -0.0785 | 0.0130001 | 1.56E-09 | 0 | SLC2A5 |
| 1 | 64948270 | rs12046291 | A | G | 0.0941 | 0.0143 | 5.08E-11 | 0.0941 | 0.0143002 | 4.69E-11 | 0 | JAK1 |
| 1 | 77501822 | rs71658797 | A | T | 0.1228 | 0.0214 | 9.83E-09 | 0.1228 | 0.0214002 | 9.57E-09 | 0 | AK5 |
| 1 | 155066988 | rs114301457 | T | C | 0.874 | 0.1446 | 1.51E-09 | 0.891334 | 0.144648 | 7.18E-10 | -0.012 | EFNA4 |
| 1 | 155175305 | rs7528026 | A | G | 0.2997 | 0.0375 | 1.32E-15 | 0.305034 | 0.0377231 | 6.16E-16 | 0.062 | TRIM46—KRTCAP2 |
| 1 | 155197995 | rs41264915 | A | G | 0.1915 | 0.0201 | 1.94E-21 | 0.21595 | 0.020708 | 1.84E-25 | 0.25 | THBS3 |
| 1 | 155278322 | rs11264349 | A | T | -0.0587 | 0.0148 | 7.29E-05 | -0.110626 | 0.0153001 | 4.82E-13 | 0 | HCN3 |
| 2 | 60480453 | rs1123573 | A | G | 0.1116 | 0.0144 | 1.01E-14 | 0.111051 | 0.0144005 | 1.24E-14 | 0 | BCL11A |
| 3 | 45796521 | rs2271616 | T | G | 0.1678 | 0.0202 | 1.10E-16 | 0.235271 | 0.0202924 | 4.42E-31 | -0.094 | SLC6A20 |
| 3 | 45818159 | rs17713054 | A | G | 0.7212 | 0.0217 | 7.64E-243 | 0.671641 | 0.023844 | 1.44E-174 | -0.72 | LZTFL1 |
| 3 | 45873093 | rs35482426 | CTT | C | -0.6303 | 0.0312 | 6.12E-91 | -0.257237 | 0.0341932 | 5.35E-14 | 0 | LZTFL1 |
| 3 | 101737122 | rs11706986 | T | C | 0.0749 | 0.0131 | 1.10E-08 | 0.0749 | 0.0131001 | 1.08E-08 | 0 | CEP97 |
| 3 | 146517122 | rs343320 | A | G | 0.1536 | 0.0255 | 1.81E-09 | 0.1536 | 0.0255003 | 1.71E-09 | 0 | PLSCR1 |
| 4 | 25446871 | rs7664615 | A | G | 0.0985 | 0.0174 | 1.46E-08 | 0.0985 | 0.0174002 | 1.51E-08 | 0 | ANAPC4 |
| 4 | 167824478 | rs1073165 | A | G | 0.0725 | 0.0129 | 1.74E-08 | 0.0725 | 0.0129001 | 1.91E-08 | 0 | . |
| 5 | 132441275 | rs10066378 | T | C | -0.1129 | 0.0176 | 1.33E-10 | -0.1129 | 0.0176002 | 1.41E-10 | 0 | IRF1—IRF1-AS1 |
| 6 | 31153455 | rs111837807 | T | C | -0.1833 | 0.0217 | 3.25E-17 | -0.221112 | 0.0220419 | 1.11E-23 | -0.15 | CCHCR1 |
| 6 | 31513129 | rs2516401 | A | G | -0.0828 | 0.0138 | 2.23E-09 | -0.0985512 | 0.0139528 | 1.63E-12 | -0.052 | MICB |
| 6 | 32702531 | rs2856717 | A | G | 0.0818 | 0.0137 | 2.16E-09 | 0.0914593 | 0.0137875 | 3.28E-11 | 0.0066 | HLA-DQA1 |
| 6 | 41522644 | rs41435745 | C | G | 0.3423 | 0.0368 | 1.50E-20 | 0.341197 | 0.0368024 | 1.84E-20 | 0 | FOXP4 |
| 7 | 75623396 | rs1179620 | T | C | -0.0824 | 0.0138 | 2.26E-09 | -0.0824 | 0.0138002 | 2.36E-09 | 0 | HIP1 |
| 7 | 100032719 | rs2897075 | T | C | 0.0786 | 0.0129 | 1.13E-09 | 0.0786 | 0.0129001 | 1.11E-09 | 0 | ZKSCAN1 |
| 8 | 60532539 | rs13276831 | T | C | 0.0703 | 0.0125 | 2.04E-08 | 0.0703 | 0.0125001 | 1.87E-08 | 0 | RAB2A |
| 9 | 21206606 | rs28368148 | C | G | -0.5284 | 0.0905 | 5.26E-09 | -0.5284 | 0.0905021 | 5.27E-09 | 0 | IFNA10 |
| 9 | 33425186 | rs60840586 | G | GTAAC | 0.0979 | 0.0171 | 9.71E-09 | 0.0979 | 0.0171002 | 1.03E-08 | 0 | AQP3 |
| 9 | 133271182 | rs879055593 | T | C | 0.1226 | 0.0148 | 1.03E-16 | 0.1226 | 0.0148003 | 1.19E-16 | 0 | ABO |
| 10 | 79946568 | rs721917 | A | G | -0.074 | 0.0127 | 5.13E-09 | -0.074 | 0.0127001 | 5.65E-09 | 0 | SFTPD |
| 11 | 1219991 | rs35705950 | T | G | -0.1583 | 0.021 | 5.52E-14 | -0.1583 | 0.0210003 | 4.77E-14 | 0 | MUC5B |
| 11 | 34482745 | rs61882275 | A | G | -0.1221 | 0.013 | 6.84E-21 | -0.1221 | 0.0130003 | 5.88E-21 | 0 | ELF5 |
| 12 | 112919637 | rs2660 | A | G | 0.096 | 0.0132 | 3.86E-13 | 0.096 | 0.0132002 | 3.53E-13 | 0 | OAS1 |
| 12 | 132481571 | rs11614702 | A | G | 0.1046 | 0.0127 | 1.52E-16 | 0.1046 | 0.0127002 | 1.78E-16 | 0 | FBRSL1 |
| 13 | 112881427 | rs12585036 | T | C | 0.1347 | 0.0151 | 3.98E-19 | 0.1347 | 0.0151003 | 4.65E-19 | 0 | ATP11A |
| 16 | 89196249 | rs117169628 | A | G | 0.1484 | 0.0184 | 7.20E-16 | 0.1484 | 0.0184003 | 7.32E-16 | 0 | SLC22A31 |
| 17 | 40003082 | rs12941811 | T | C | -0.0791 | 0.0127 | 4.46E-10 | -0.0795126 | 0.0127002 | 3.83E-10 | -0.0039 | PSMD3 |
| 17 | 46085231 | rs8080583 | A | C | -0.1253 | 0.015 | 7.99E-17 | -0.126009 | 0.0150004 | 4.45E-17 | 0.0031 | KANSL1 |
| 17 | 49863303 | rs77534576 | T | C | 0.2964 | 0.0372 | 1.51E-15 | 0.297299 | 0.0372008 | 1.33E-15 | 0 | TAC4 |
| 19 | 4717660 | rs12610495 | A | G | -0.2293 | 0.0153 | 9.06E-51 | -0.226943 | 0.015302 | 9.25E-50 | -0.0092 | DPP9 |
| 19 | 10352442 | rs34536443 | C | G | 0.4034 | 0.0365 | 2.24E-28 | 0.418348 | 0.0365594 | 2.55E-30 | -0.064 | TYK2 |
| 19 | 10414696 | rs142770866 | A | G | 0.2189 | 0.024 | 7.62E-20 | 0.23209 | 0.024038 | 4.68E-22 | 0 | PDE4A |
| 19 | 48702888 | rs516316 | C | G | -0.0924 | 0.0127 | 3.74E-13 | -0.0915819 | 0.0127007 | 5.56E-13 | -0.009 | FUT2 |
| 19 | 50374423 | rs35463555 | A | G | 0.0986 | 0.0136 | 4.16E-13 | 0.0977207 | 0.0136007 | 6.72E-13 | 0 | NR1H2 |
| 21 | 33229937 | rs188401375 | C | G | -0.2962 | 0.0635 | 3.05E-06 | -0.376012 | 0.0637017 | 3.58E-09 | -0.11 | IFNAR2 |
| 21 | 33237639 | rs9636867 | A | G | -0.1968 | 0.0134 | 8.71E-49 | -0.190179 | 0.0135242 | 6.49E-45 | -0.13 | IFNAR2 |
| 21 | 33287378 | rs8178521 | T | C | 0.1245 | 0.016 | 6.15E-15 | 0.10378 | 0.0161191 | 1.21E-10 | -0.019 | IL10RB |
| 21 | 33949755 | rs75912728 | T | C | 0.1611 | 0.0205 | 3.60E-15 | 0.157668 | 0.0205073 | 1.49E-14 | 0.0021 | ATP5PO |
| 21 | 41479527 | rs915823 | A | C | 0.0969 | 0.0161 | 1.70E-09 | 0.0943654 | 0.0161009 | 4.60E-09 | 0 | TMPRSS2 |
| 23 | 15523993 | rs35697037 | A | G | 0.0421 | 0.0073 | 6.81E-09 | 0.0421 | 0.00730002 | 8.06E-09 | 0 | ACE2—BMX |

Table 3: Condtional analysis for critical illness phenotype. Conditional analysis was calculated using EUR reference panel as most of the population from the analysis is from European ancestry. Chr: chromosome, Pos(b38): position on human genome build 38, Allele1: effect allele, Allele2: other allele, beta: effect, SE: standard error, P: P-value, $beta_{cond}$: beta in conditional analysis $SE_{cond}$: standard error in conditional analysis $P_{cond}$: P-value in conditional analysis $LD_r$: LD between $SNP_i$ and $SNP_{i+1}$ significant in conditional analysis, Gene: nearest or most plausible nearby gene

| Chr | Pos(b38) | rsid | Allele1 | Allele2 | OR | $OR_{CI}$ | P | Het P.value | N | Gene | Citation |
|---|---|---|---|---|---|---|---|---|---|---|---|
| 1 | 64947147 | rs11208552 | T | G | 0.94 | 0.93-0.96 | 2.7e-09 | 0.38 | 2818410 | JAK1 | GenOMICC *new* |
| 1 | 77501822 | rs71658797 | A | T | 1.1 | 1.06-1.13 | 4.8e-09 | 0.17 | 2940840 | AK5 | GenOMICC *new* |
| 1 | 155003661 | rs61811895 | T | G | 1.1 | 1.06-1.12 | 1.2e-10 | 0.027 | 2652420 | ZBTB7B | GenOMICC *new* |
| 1 | 155066988 | rs114301457 | T | C | 1.7 | 1.46-2.02 | 3.7e-11 | 0.0051 | 1557660 | EFNA4 | GenOMICC[7] |
| 1 | 155172916 | rs41264911 | A | G | 1.2 | 1.16-1.29 | 1.4e-13 | 0.056 | 2945320 | TRIM46 | GenOMICC[7] |
| 1 | 155197995 | rs41264915 | A | G | 1.1 | 1.11-1.17 | 1.5e-21 | 0.16 | 2685850 | THBS3 | HGI[5] |
| 2 | 60480453 | rs1123573 | A | G | 1.1 | 1.05-1.09 | 1.5e-12 | 0.0014 | 2921600 | BCL11A | GenOMICC[7] |
| 3 | 45696221 | rs149853133 | T | C | 0.7 | 0.63-0.77 | 2e-12 | 0.39 | 2499890 | SACM1L | GenOMICC *new* |
| 3 | 45796521 | rs2271616 | T | G | 1.1 | 1.11-1.17 | 4.8e-20 | 7e-06 | 2090810 | SLC6A20 | HGI[5] |
| 3 | 45818159 | rs17713054‡ | A | G | 1.7 | 1.61-1.72 | 1.5e-229 | 4.4e-40 | 2946110 | LZTFL1 | SCGG[8] |
| 3 | 101778725 | rs1460097 | T | C | 0.94 | 0.92-0.95 | 2.9e-12 | 0.23 | 2952250 | NXPE3 | GenOMICC *new* |
| 3 | 146514682 | rs454645 | T | C | 1.1 | 1.08-1.15 | 7.5e-12 | 0.076 | 2942040 | PLSCR1 | GenOMICC[7] |
| 4 | 25447603 | rs7671107 | A | G | 0.93 | 0.91-0.95 | 9.6e-10 | 0.093 | 2876850 | ARHGEF38 | GenOMICC *new* |
| 5 | 132075767 | rs25882 | T | C | 0.94 | 0.92-0.96 | 1.9e-10 | 0.0097 | 2960520 | CSF2 | (GenOMICC[7]) |
| 6 | 31153649 | rs143334143‡ | A | G | 1.1 | 1.11-1.17 | 4.2e-17 | 1.5e-11 | 2947720 | CCHCR1 | GenOMICC[2] |
| 6 | 31306250 | rs1634761 | T | C | 0.94 | 0.92-0.96 | 1.1e-12 | 0.49 | 2961060 | HLA-B | GenOMICC *new* |
| 6 | 41522644 | rs41435745 | C | G | 1.2 | 1.18-1.29 | 1.2e-20 | 0.019 | 2834490 | FOXP4 | HGI[5] |
| 7 | 100032719 | rs2897075 | T | C | 1.1 | 1.03-1.07 | 3.4e-08 | 0.38 | 2959610 | ZKSCAN1 | GenOMICC *new* |
| 8 | 60507169 | rs2875974 | A | G | 1.1 | 1.04-1.07 | 5.8e-10 | 0.23 | 2960030 | RAB2A | GenOMICC *new* |
| 9 | 15795835 | rs79611697 | T | G | 1.1 | 1.08-1.16 | 4.1e-08 | 0.061 | 2942740 | CCDC171 | GenOMICC *new* |
| 9 | 21206606 | rs28368148 | C | G | 0.75 | 0.68-0.83 | 2.7e-08 | 0.014 | 2326800 | IFNA10 | GenOMICC[2] |
| 9 | 33425787 | rs11790730 | T | C | 0.93 | 0.91-0.95 | 2.7e-09 | 0.062 | 2789950 | AQP3 | SCOURGE[4] |
| 9 | 133257521 | rs77641731 | T | TC | 0.9 | 0.89-0.92 | 2.4e-23 | 0.2 | 1975010 | ABO | SCGG[8] |
| 10 | 79946568 | rs721917 | A | G | 0.95 | 0.93-0.96 | 1.9e-10 | 0.18 | 2961130 | SFTPD | HGI[9] |
| 11 | 1219991 | rs35705950 | T | G | 0.9 | 0.87-0.92 | 5.2e-14 | 0.36 | 2664500 | MUC5B | HGI[9] |
| 11 | 34480495 | rs7949972 | T | C | 0.91 | 0.89-0.92 | 3.3e-27 | 0.028 | 2952950 | ELF5 | GenOMICC[7] |
| 12 | 112919637 | rs2660 | A | G | 1.1 | 1.06-1.1 | 8.1e-18 | 0.19 | 2937630 | OAS1 | GenOMICC[2] |
| 12 | 132565387 | rs5023077 | T | C | 1.1 | 1.05-1.09 | 1.2e-13 | 0.24 | 2829560 | FBRSL1 | GenOMICC[7] |
| 13 | 112881427 | rs12585036 | T | C | 1.1 | 1.08-1.13 | 9e-20 | 0.0078 | 2734490 | ATP11A | GenOMICC[7] |
| 16 | 54221310 | rs2102497 | T | C | 0.94 | 0.92-0.96 | 2.3e-08 | 0.78 | 2149180 | IRX3 | GenOMICC *new* |
| 16 | 89196249 | rs117169628 | A | G | 1.1 | 1.09-1.15 | 2.6e-17 | 0.084 | 2952150 | SLC22A31 | GenOMICC[7] |
| 17 | 45983409 | rs63750417 | T | C | 0.9 | 0.89-0.92 | 7.2e-19 | 0.34 | 2938130 | MAPT | GenOMICC *new* |
| 19 | 4063488 | rs66833742 | T | C | 0.93 | 0.91-0.95 | 2.1e-10 | 0.082 | 2918950 | ZBTB7A | GenOMICC *new* |
| 19 | 4717660 | rs12610495‡ | A | G | 0.86 | 0.84-0.88 | 1.4e-48 | 3.2e-09 | 2887040 | DPP9 | GenOMICC[2] |
| 19 | 10364976 | rs2304256 | A | C | 1.1 | 1.08-1.12 | 1.2e-23 | 0.019 | 2961600 | TYK2 | GenOMICC[2] |
| 19 | 48702851 | rs679574 | C | G | 1.1 | 1.04-1.08 | 6.4e-11 | 0.058 | 2949460 | FUT2 | GenOMICC[7] |
| 19 | 50379362 | rs1405655 | T | C | 0.93 | 0.91-0.94 | 2.1e-16 | 0.21 | 2961590 | NR1H2 | HGI[9] |
| 20 | 6517380 | rs6117308 | A | G | 0.95 | 0.93-0.97 | 9.1e-09 | 0.71 | 2961590 | CASC20 | GenOMICC *new* |
| 21 | 33237639 | rs9636867 | A | G | 0.87 | 0.85-0.89 | 2e-50 | 0.00042 | 2961120 | IFNAR2 | GenOMICC[2] |
| 21 | 33934844 | rs78143111 | A | G | 1.1 | 1.11-1.17 | 3.6e-17 | 0.0054 | 2953580 | ATP5PO | GenOMICC[7] |
| 21 | 41479527 | rs915823 | A | C | 1.1 | 1.06-1.1 | 1.8e-11 | 0.81 | 2954430 | TMPRSS2 | GenOMICC *new* |
| 23 | 15497196 | | C | G | 0.96 | 0.95-0.97 | 2.9e-11 | 0.45 | 2377330 | ACE2 | HGI[9] |

Table 4: Full results for severe (hospitalised) covid GWAS. Chr: chromosome, Pos(b38): position on human genome build 38, Allele1: effect allele, Allele2: other allele, OR: odds ratio, $OR_{CI}$: confidence interval, N: number of individuals in the analysis Het P.value: Heterogeneity P-value Gene: nearest or most plausible nearby gene, Citation: first demonstration of association. ‡ indicates significant heterogeneity across studies.

**GenOMICC Consortium** J. Kenneth Baillie[2,3], Colin Begg[4], Sara Clohisey[2], Charles Hinds[5], Peter Horby[6], Julian Knight[7], Lowell Ling[8], David Maslove[9], Danny McAuley[10,11], Johnny Millar[2], Hugh Montgomery[12], Alistair Nichol[13], Peter J.M. Openshaw[14,15], Alexandre C Pereira[16], Chris P Ponting[17], Kathy Rowan[18], Malcolm G Semple[19,20], Manu Shankar-Hari[21], Charlotte Summers[22], Timothy Walsh[3], Latha Aravindan[23], Ruth Armstrong[2], J. Kenneth Baillie[2,3], Heather Biggs[24], Ceilia Boz[2], Adam Brown[2], Richard Clark[25], Sara Clohisey[2], Audrey Coutts[25], Judy Coyle[2], Louise Cullum[2], Sukamal Das[23], Nicky Day[2], Lorna Donnelly[25], Esther Duncan[2], Angie Fawkes[25], Paul Fineran[2], Max Head Fourman[2], Anita Furlong[24], James Furniss[2], Bernadette Gallagher[2], Tammy Gilchrist[25], Ailsa Golightly[2], Fiona Griffiths[2], Katarzyna Hafezi[25], Debbie Hamilton[2], Ross Hendry[2], Andy Law[2], Dawn Law[2], Rachel Law[2], Sarah Law[2], Rebecca Lidstone-Scott[2], Louise Macgillivray[25], Alan Maclean[25], Hanning Mal[2], Sarah McCafferty[25], Ellie Mcmaster[2], Jen Meikle[2], Shona C Moore[19], Kirstie Morrice[25], Lee Murphy[25], Sheena Murphy[23], Mybaya Hellen[2], Wilna Oosthuyzen[2], Chenqing Zheng[26], Jiantao Chen[26], Nick Parkinson[2], Trevor Paterson[2], Katherine Schon[24], Andrew Stenhouse[2], Mihaela Das[23], Maaike Swets[2,27], Helen Szoor-McElhinney[2], Filip Taneski[2], Lance Turtle[19], Tony Wackett[2], Mairi Ward[2], Jane Weaver[2], Nicola Wrobel[25], Marie Zechner[2], Mybaya Hellen[2], Gill Arbane[28], Aneta Bociek[28], Sara Campos[28], Neus Grau[28], Tim Owen Jones[28], Rosario Lim[28], Martina Marotti[28], Marlies Ostermann[28], Manu Shankar-Hari[28], Christopher Whitton[28], Zoe Alldis[29], Raine Astin-Chamberlain[29], Fatima Bibi[29], Jack Biddle[29], Sarah Blow[29], Matthew Bolton[29], Catherine Borra[29], Ruth Bowles[29], Maudrian Burton[29], Yasmin Choudhury[29], David Collier[29], Amber Cox[29], Amy Easthope[29], Patrizia Ebano[29], Stavros Fotiadis[29], Jana Gurasashvili[29], Rosslyn Halls[29], Pippa Hartridge[29], Delordson Kallon[29], Jamila Kassam[29], Ivone Lancoma-Malcolm[29], Maninderpal Matharu[29], Peter May[29], Oliver Mitchelmore[29], Tabitha Newman[29], Mital Patel[29], Jane Pheby[29], Irene Pinzuti[29], Zoe Prime[29], Oleksandra Prysyazhna[29], Julian Shiel[29], Melanie Taylor[29], Carey Tierney[29], Suzanne Wood[29], Anne Zak[29], Olivier Zongo[29], Stephen Bonner[30], Keith Hugill[30], Jessica Jones[30], Steven Liggett[30], Evie Headlam[30], Nageswar Bandla[31], Minnie Gellamucho[31], Michelle Davies[31], Christopher Thompson[31], Marwa Abdelrazik[32], Dhanalakshmi Bakthavatsalam[32], Munzir Elhassan[32], Arunkumar Ganesan[32], Anne Haldeos[32], Jeronimo Moreno-Cuesta[32], Dharam Purohit[32], Rachel Vincent[32], Kugan Xavier[32], kumar Rohit[33], Frater Alasdair[32], Malik Saleem[32], Carter David[32], Jenkins Samuel[32], Zoe Lamond[32], Wall Alanna[32], Jaime Fernandez-Roman[34], David O. Hamilton[34], Emily Johnson[34], Brian Johnston[34], Maria Lopez Martinez[34], Suleman Mulla[34], David Shaw[34], Alicia A.C. Waite[34], Victoria Waugh[34], Ingeborg D. Welters[34], Karen Williams[34], Anna Cavazza[35], Maeve Cockrell[35], Eleanor Corcoran[35], Maria Depante[35], Clare Finney[35], Ellen Jerome[35], Mark McPhail[35], Monalisa Nayak[35], Harriet Noble[35], Kevin O'Reilly[35], Evita Pappa[35], Rohit Saha[35], Sian Saha[35], John Smith[35], Abigail Knighton[35], David Antcliffe[36], Dorota Banach[36], Stephen Brett[36], Phoebe Coghlan[36], Ziortza Fernandez[36], Anthony Gordon[36], Roceld Rojo[36], Sonia Sousa Arias[36], Maie Templeton[36], Megan Meredith[37], Lucy Morris[37], Lucy Ryan[37], Amy Clark[37], Julia Sampson[37], Cecilia Peters[37], Martin Dent[37], Margaret Langley[37], Saima Ashraf[37], Shuying Wei[37], Angela Andrew[37], Archana Bashyal[38], Neil Davidson[38], Paula Hutton[38], Stuart McKechnie[38], Jean Wilson[38], David Baptista[39], Rebecca Crowe[39], Rita Fernandes[39], Rosaleen Herdman-Grant[39], Anna Joseph[39], Denise O'Connor[40], Meryem Allen[39], Adam Loveridge[39], India McKenley[39], Eriko Morino[39], Andres Naranjo[39], Richard Simms[39], Kathryn Sollesta[39], Andrew Swain[39], Harish Venkatesh[39], Jacyntha Khera[39], Jonathan Fox[39], Gillian Andrew[41], J. Kenneth Baillie[41], Lucy Barclay[41], Marie Callaghan[41], Rachael Campbell[41], Sarah Clark[41], Dave Hope[41], Lucy Marshall[41], Corrienne McCulloch[41], Kate Briton[41], Jo Singleton[41], Sohphie Birch[41], Lutece Brimfield[42], Zoe Daly[42], David Pogson[42], Steve Rose[42], Ceri Battle[43], Elaine Brinkworth[43], Rachel Harford[43], Carl Murphy[43], Luke Newey[43], Tabitha Rees[43], Marie Williams[43], Sophie Arnold[43], Petra Polgarova[44], Katerina Stroud[44], Charlotte Summers[44], Eoghan Meaney[44], Megan Jones[44], Anthony Ng[44], Shruti Agrawal[44], Nazima Pathan[44], Deborah White[44], Esther Daubney[44], Kay Elston[44], Lina Grauslyte[45], Musarat Hussain[45], Mandeep Phull[45], Tatiana Pogreban[45], Lace Rosaroso[45], Erika Salciute[45], George Franke[45], Joanna Wong[45], Aparna George[45], Laura Ortiz-Ruiz de Gordoa[46], Emily Peasgood[46], Claire Phillips[46], Laura Ortiz-Ruiz de Gordoa[46], Emily Peasgood[46], Claire Phillips[46], Michelle Bates[47], Jo Dasgin[47], Jaspret Gill[47], Annette Nilsson[47], James Scriven[47], Carlos Castro Delgado[48], Deborah Dawson[48], Lijun Ding[48], Georgia Durrant[48], Obiageri Ezeobu[48], Sarah Farnell-Ward[48], Abiola Harrison[48], Rebecca Kanu[48], Susannah Leaver[48], elena Maccacari[48], Soumendu Manna[48], Romina Pepermans Saluzzio[48], Joana Queiroz[48], Tinashe Samakomva[48], Christine Sicat[48], Joana Texeira[48], Edna Fernandes Da Gloria[48], Ana Lisboa[48], John Rawlins[48], Jisha Mathew[48], Ashley Kinch[48], William James Hurt[48], Nirav Shah[48], Victoria Clark[48], Maria Thanasi[48], Nikki Yun[48], Kamal

Patel[48], Sara Bennett[49], Emma Goodwin[49], Matthew Jackson[49], Alissa Kent[49], Clare Tibke[49], Wiesia Woodyatt[49], Ahmed Zaki[49], Azmerelda Abraheem[50], Peter Bamford[50], Kathryn Cawley[50], Charlie Dunmore[50], Maria Faulkner[50], Rumanah Girach[50], Helen Jeffrey[50], Rhianna Jones[50], Emily London[50], Imrun Nagra[50], Farah Nasir[50], Hannah Sainsbury[50], Clare Smedley[50], Tahera Patel[51], Matthew Smith[51], Srikanth Chukkambotla[51], Aayesha Kazi[51], Janice Hartley[51], Joseph Dykes[51], Muhammad Hijazi[51], Sarah Keith[51], Meherunnisa Khan[51], Janet Ryan-Smith[51], Philippa Springle[51], Jacqueline Thomas[51], Nick Truman[51], Samuel Saad[51], Dabheoc Coleman[51], Christopher Fine[51], Roseanna Matt[51], Bethan Gay[51], Jack Dalziel[51], Syamlan Ali[51], Drew Goodchild[51], Rhiannan Harling[51], Ravi Bhatterjee[51], Wendy Goddard[51], Chloe Davison[51], Stephen Duberly[51], Jeanette Hargreaves[51], Rachel Bolton[51], Miriam Davey[52], David Golden[52], Rebecca Seaman[52], Shiney Cherian[53], Sean Cutler[53], Anne Emma Heron[53], Anna Roynon-Reed[53], Tamas Szakmany[53], Gemma Williams[53], Owen Richards[53], Yusuf Cheema[53], Hollie Brooke[54], Sarah Buckley[54], Jose Cebrian Suarez[54], Ruth Charlesworth[54], Karen Hansson[54], John Norris[54], Alice Poole[54], Alastair Rose[54], Rajdeep Sandhu[54], Brendan Sloan[54], Elizabeth Smithson[54], Muthu Thirumaran[54], Veronica Wagstaff[54], Alexandra Metcalfe[54], Mark Brunton[55], Jess Caterson[55], Holly Coles[55], Matthew Frise[55], Sabi Gurung Rai[55], Nicola Jacques[55], Liza Keating[55], Emma Tilney[55], Shauna Bartley[55], Parminder Bhuie[55], Sian Gibson[56], Amanda Lyle[56], Fiona McNeela[56], Jayachandran Radhakrishnan[56], Alistair Hughes[56], Bryan Yates[57], Jessica Reynolds[57], Helen Campbell[57], Maria Thompsom[57], Steve Dodds[57], Stacey Duffy[57], Sandra Greer[58], Karen Shuker[58], Ascanio Tridente[58], Reena Khade[59], Ashok Sundar[59], George Tsinaslanidis[59], Isobel Birkinshaw[60], Joseph Carter[60], Kate Howard[60], Joanne Ingham[60], Rosie Joy[60], Harriet Pearson[60], Samantha Roche[60], Zoe Scott[60], Hollie Bancroft[61], Mary Bellamy[61], Margaret Carmody[61], Jacqueline Daglish[61], Faye Moore[61], Joanne Rhodes[61], Mirriam Sangombe[61], Salma Kadiri[61], James Scriven[61], Maria Croft[62], Ian White[62], Victoria Frost[62], Maia Aquino[62], Rajeev Jha[63], Vinodh Krishnamurthy[63], Lai Lim[63], Rajeev Jha[63], Vinodh Krishnamurthy[63], Li Lim[63], Edward Combes[64], Teishel Joefield[64], Sonja Monnery[64], Valerie Beech[64], Sallyanne Trotman[64], Christine Almaden-Boyle[65], Pauline Austin[65], Louise Cabrelli[65], Stephen Cole[65], Matt Casey[65], Susan Chapman[65], Stephen Cole[65], Clare Whyte[65], Yolanda Baird[66], Aaron Butler[66], Indra Chadbourn[66], Linda Folkes[66], Heather Fox[66], Amy Gardner[66], Raquel Gomez[66], Gillian Hobden[66], Luke Hodgson[66], Kirsten King[66], Michael Margarson[66], Tim Martindale[66], Emma Meadows[66], Dana Raynard[66], Yvette Thirlwall[66], David Helm[66], Jordi Margalef[66], Kristine Criste[67], Rebecca Cusack[67], Kim Golder[67], Hannah Golding[67], Oliver Jones[67], Samantha Leggett[67], Michelle Male[67], Martyna Marani[67], Kirsty Prager[67], Toran Williams[67], Belinda Roberts[67], Karen Salmon[67], Peter Anderson[68], Katie Archer[68], Karen Austin[68], caroline Davis[68], Alison Durie[68], Olivia Kelsall[68], Jessica Thrush[68], Charlie Vigurs[68], Laura Wild[68], Hannah-Louise Wood[68], Helen Tranter[68], Alison Harrison[68], Nicholas Cowley[68], Michael McAlindon[68], Andrew Burtenshaw[68], Stephen Digby[68], Emma Low[68], Aled Morgan[68], Naiara Cother[68], Tobias Rankin[68], Sarah Clayton[68], Alex McCurdy[68], Cecilia Ahmed[69], Balvinder Baines[69], Sarah Clamp[69], Julie Colley[69], Risna Haq[69], Anne Hayes[69], Jonathan Hulme[69], Samia Hussain[69], Sibet Joseph[69], Rita Kumar[69], Zahira Maqsood[69], Manjit Purewal[69], Leonie Benham[70], Zena Bradshaw[70], Joanna Brown[70], Melanie Caswell[70], Jason Cupitt[70], Sarah Melling[70], Stephen Preston[70], Nicola Slawson[70], Emma Stoddard[70], Scott Warden[70], Bethan Deacon[71], Ceri Lynch[71], Carla Pothecary[71], Lisa Roche[71], Gwenllian Sera Howe[71], Jayaprakash Singh[71], Keri Turner[71], Hannah Ellis[71], Natalie Stroud[71], Jodie Hunt[72], Joy Dearden[72], Emma Dobson[72], Andy Drummond[72], Michelle Mulcahy[72], Sheila Munt[72], Grainne O'Connor[72], Jennifer Philbin[72], Chloe Rishton[72], Redmond Tully[72], Sarah Winnard[72], Susanne Cathcart[73], Katharine Duffy[73], Alex Puxty[73], Kathryn Puxty[73], Lynne Turner[73], Jane Ireland[73], Gary Semple[73], Kate Long[74], Simon Whiteley[74], Elizabeth Wilby[74], Bethan Ogg[74], Amanda Cowton[75], Andrea Kay[75], Melanie Kent[75], Kathryn Potts[75], Ami Wilkinson[75], Suzanne Campbell[75], Ellen Brown[75], Julie Melville[76], Jay Naisbitt[76], Rosane Joseph[76], Maria Lazo[76], Olivia Walton[76], Alan Neal[76], Peter Alexander[77], Schvearn Allen[77], Joanne Bradley-Potts[77], Craig Brantwood[77], Jasmine Egan[77], Timothy Felton[77], Grace Padden[77], Luke Ward[77], Stuart Moss[77], Susannah Glasgow[77], Lynn Abel[78], Michael Brett[78], Brian Digby[78], Lisa Gemmell[78], James Hornsby[78], Patrick MacGoey[78], Pauline O'Neil[78], Richard Price[78], Natalie Rodden[78], Kevin Rooney[78], Radha Sundaram[78], Nicola Thomson[78], Bridget Hopkins[79], James Scriven[79], Laura Thrasyvoulou[79], Heather Willis[79], Martyn Clark[80], Martina Coulding[80], Edward Jude[80], Jacqueline McCormick[80], Oliver Mercer[80], Darsh Potla[80], Hafiz Rehman[80], Heather Savill[80], Victoria Turner[80], Charlotte Downes[81], Kathleen Holding[81], Katie Riches[81], Mary Hilton[81], Mel Hayman[81], Deepak Subramanian[81], Priya Daniel[81], Oluronke Adanini[82], Nikhil Bhatia[82], Maines Msiska[82], Rebecca Collins[82], Ian Clement[83], Bijal Patel[83], A Gulati[83], Carole Hays[83], K Webster[83], Anne Hudson[83], Andrea Webster[83], Elaine

Stephenson[83], Louise McCormack[83], Victoria Slater[83], Rachel Nixon[83], Helen Hanson[83], Maggie fearby[83], Sinead Kelly[83], Victoria Bridgett[83], Philip Robinson[83], Julie Camsooksai[84], Charlotte Humphrey[84], Sarah Jenkins[84], Henrik Reschreiter[84], Beverley Wadams[84], Yasmin Death[84], Victoria Bastion[85], Daphene Clarke[85], Beena David[85], Harriet Kent[85], Rachel Lorusso[85], Gamu Lubimbi[85], Sophie Murdoch[85], Melchizedek Penacerrada[85], Alastair Thomas[85], Jennifer Valentine[85], Ana Vochin[85], Retno Wulandari[85], Brice Djeugam[85], Gillian Bell[86], Katy English[86], Amro Katary[86], Louise Wilcox[86], Michelle Bruce[87], Karen Connolly[87], Tracy Duncan[87], Helen T-Michael[87], Gabriella Lindergard[87], Samuel Hey[87], Claire Fox[87], Jordan Alfonso[87], Laura Jayne Durrans[87], Jacinta Guerin[87], Bethan Blackledge[87], Jade Harris[87], Martin Hruska[87], Ayaa Eltayeb[87], Thomas Lamb[87], Tracey Hodgkiss[87], Lisa Cooper[87], Joanne Rothwell[87], Angela Allan[88], Felicity Anderson[88], Callum Kaye[88], Jade Liew[88], Jasmine Medhora[88], Teresa Scott[88], Erin Trumper[88], Adriana Botello[88], Liana Lankester[89], Nikitas Nikitas[89], Colin Wells[89], Bethan Stowe[89], Kayleigh Spencer[89], Craig Brandwood[90], Lara Smith[90], Richard Clark[90], Katie Birchall[90], Laurel Kolakaluri[90], Deborah Baines[90], Anila Sukumaran[90], Elena Apetri[91], Cathrine Basikolo[91], Bethan Blackledge[91], Laura Catlow[91], Bethan Charles[91], Paul Dark[91], Reece Doonan[91], Jade Harris[91], Alice Harvey[91], Daniel Horner[91], Karen Knowles[91], Stephanie Lee[91], Diane Lomas[91], Chloe Lyons[91], Tracy Marsden[91], Danielle McLaughlan[91], Liam McMorrow[91], Jessica Pendlebury[91], Jane Perez[91], Maria Poulaka[91], Nicola Proudfoot[91], Melanie Slaughter[91], Kathryn Slevin[91], Melanie Taylor[91], Vicky Thomas[91], Danielle Walker[91], Angiy Michael[91], Matthew Collis[91], Tracey Cosier[92], Gemma Millen[92], Neil Richardson[92], Natasha Schumacher[92], Heather Weston[92], James Rand[92], Nicola Baxter[93], Steven Henderson[93], Sophie Kennedy-Hay[93], Christopher McParland[93], Laura Rooney[93], Malcolm Sim[93], Gordan McCreath[93], Louise Akeroyd[94], Shereen Bano[94], Matt Bromley[94], Lucy Gurr[94], Tom Lawton[94], James Morgan[94], Kirsten Sellick[94], Deborah Warren[94], Brian Wilkinson[94], Janet McGowan[94], Camilla Ledgard[94], Amelia Stacey[94], Kate Pye[94], Ruth Bellwood[94], Michael Bentley[94], Jeremy Bewley[95], Zoe Garland[95], Lisa Grimmer[95], Bethany Gumbrill[95], Rebekah Johnson[95], Katie Sweet[95], Denise Webster[95], Georgia Efford[95], Karen Convery[96], Deirdre Fottrell-Gould[96], Lisa Hudig[96], Jocelyn Keshet-Price[96], Georgina Randell[96], Katie Stammers[96], Maria Bokhari[97], Vanessa Linnett[97], Rachael Lucas[97], Wendy McCormick[97], Jenny Ritzema[97], Amanda Sanderson[97], Helen Wild[97], Anthony Rostron[98], Alistair Roy[98], Lindsey Woods[98], Sarah Cornell[98], Fiona Wakinshaw[98], Kimberley Rogerson[98], Jordan Jarmain[98], Robert Parker[99], Amie Reddy[99], Ian Turner-Bone[99], Laura Wilding[99], Peter Harding[99], Caroline Abernathy[100], Louise Foster[100], Andrew Gratrix[100], Vicky Martinson[100], Priyai Parkinson[100], Elizabeth Stones[100], Llucia Carbral-Ortega[101], Georgia Bercades[102], David Brealey[102], Ingrid Hass[102], Niall MacCallum[102], Gladys Martir[102], Eamon Raith[102], Anna Reyes[102], Deborah Smyth[102], Letizia Zitter[103], Sarah Benyon[103], Suzie Marriott[103], Linda Park[103], Samantha Keenan[103], Elizabeth Gordon[103], Helen Quinn[103], Kizzy Baines[103], Lenka Cagova[104], Adama Fofano[104], Lucie Garner[104], Helen Holcombe[104], Sue Mepham[104], Alice Michael Mitchell[104], Lucy Mwaura[104], Krithivasan Praman[104], Alain Vuylsteke[104], Julie Zamikula[104], Bally Purewal[105], Vanessa Rivers[105], Stephanie Bell[105], Hayley Blakemore[106], Borislava Borislavova[106], Beverley Faulkner[106], Emma Gendall[106], Elizabeth Goff[106], Kati Hayes[106], Matt Thomas[106], Ruth Worner[106], Kerry Smith[106], Deanna Stephens[106], Louise Mew[107], Esther Mwaura[107], Richard Stewart[107], Felicity Williams[107], Lynn Wren[107], Sara-Beth Sutherland[107], Emily Bevan[108], Jane Martin[108], Dawn Trodd[108], Geoff Watson[108], Caroline Wrey Brown[108], Amy Collins[109], Waqas Khaliq[109], Estefania Treus Gude[109], Olugbenga Akinkugbe[110], Alasdair Bamford[110], Emily Beech[110], Holly Belfield[110], Michael Bell[110], Charlene Davies[110], Gareth A. L. Jones[110], Tara McHugh[110], Hamza Meghari[110], Lauran O'Neill[110], Mark J. Peters[110], Samiran Ray[110], Ana Luisa Tomas[110], Iona Burn[111], Geraldine Hambrook[111], Katarina Manso[111], Ruth Penn[111], Pradeep Shanmugasundaram[111], Julie Tebbutt[111], Danielle Thornton[111], Jade Cole[112], Michelle Davies[112], Rhys Davies[112], Donna Duffin[112], Helen Hill[112], Ben Player[112], Emma Thomas[112], Angharad Williams[112], Denise Griffin[113], Nycola Muchenje[113], Mcdonald Mupudzi[113], Richard Partridge[113], Jo-Anna Conyngham[113], Rachel Thomas[113], Mary Wright[113], Maria Alvarez Corral[113], Reni Jacob[114], Cathy Jones[114], Craig Denmade[114], Sarah Beavis[115], Katie Dale[115], Rachel Gascoyne[115], Joanne Hawes[115], Kelly Pritchard[115], Lesley Stevenson[115], Amanda Whileman[115], Patricia Doble[116], Joanne Hutter[116], corinne Pawley[116], Charmaine Shovelton[116], Marius Vaida[116], Deborah Butcher[117], Susie O'Sullivan[117], Nicola Butterworth-Cowin[117], Norfaizan Ahmad[118], Joann Barker[118], Kris Bauchmuller[118], Sarah Bird[118], Kay Cawthron[118], Kate Harrington[118], Yvonne Jackson[118], Faith Kibutu[118], Becky Lenagh[118], Shamiso Masuko[118], Gary H Mills[118], Ajay Raithatha[118], Matthew Wiles[118], Jayne Willson[118], Helen Newell[118], Alison Lye[118], Lorenza Nwafor[118], Claire Jarman[118], Sarah Rowland-Jones[118], David Foote[118], Joby Cole[118], Roger Thompson[118], James Watson[118], Lisa Hesseldon[118], Irene Macharia[118], Luke Chetam[118], Jacqui

Smith[118], Amber Ford[118], Samantha Anderson[118], Kathryn Birchall[118], Kay Housley[118], Sara Walker[118], Leanne Milner[118], Helena Hanratty[118], Helen Trower[118], Patrick Phillips[118], Simon Oxspring[118], Ben Donne[118], Catherine Jardine[119], Dewi Williams[119], Alasdair Hay[119], Rebecca Flanagan[120], Gareth Hughes[120], scott Latham[120], Emma McKenna[120], Jennifer Anderson[120], Robert Hull[120], Kat Rhead[120], Carina Cruz[121], Natalie Pattison[121], Rob Charnock[122], Denise McFarland[122], Denise Cosgrove[122], Ashar Ahmed[123], Anna Morris[123], Srinivas Jakkula[123], Asifa Ali[124], Megan Brady[124], Sam Dale[124], Annalisa Dance[124], Lisa Gledhill[124], Jill Greig[124], Kathryn Hanson[124], Kelly Holdroyd[124], Marie Home[124], Diane Kelly[124], Ross Kitson[124], Lear Matapure[124], Deborah Melia[124], Samantha Mellor[124], Tonicha Nortcliffe[124], Jez Pinnell[124], Matthew Robinson[124], Lisa Shaw[124], Ryan Shaw[124], Lesley Thomis[124], Alison Wilson[124], Tracy Wood[124], Lee-Ann Bayo[124], Ekta Merwaha[124], Tahira Ishaq[124], Sarah Hanley[124], Bethan Deacon[125], Meg Hibbert[125], Carla Pothecary[125], Dariusz Tetla[125], Chrsitopher Woodford[125], Latha Durga[125], Gareth Kennard-Holden[125], Debbie Branney[126], Jordan Frankham[126], Sally Pitts[126], Nigel White[126], Shondipon Laha[127], Mark Verlander[127], Alexandra Williams[127], Abdelhakim Altabaibeh[128], Ana Alvaro[128], Kayleigh Gilbert[128], Louise Ma[128], Loreta Mostoles[128], Chetan Parmar[128], Kathryn Simpson[128], Champa Jetha[128], Lauren Booker[128], Anezka Pratley[128], Colene Adams[129], Anita Agasou[129], Tracie Arden[129], Amy Bowes[129], Pauline Boyle[129], Mandy Beekes[129], Heather Button[129], Nigel Capps[129], Mandy Carnahan[129], Anne Carter[129], Danielle Childs[129], Denise Donaldson[129], Kelly Hard[129], Fran Hurford[129], Yasmin Hussain[129], Ayesha Javaid[129], James Jones[129], Sanal Jose[129], Michael Leigh[129], Terry Martin[129], Helen Millward[129], Nichola Motherwell[129], Rachel Rikunenko[129], Jo Stickley[129], Julie Summers[129], Louise Ting[129], Helen Tivenan[129], Louise Tonks[129], Rebecca Wilcox[129], Maureen Holland[130], Natalie Keenan[130], Marc Lyons[130], Helen Wassall[130], Chris Marsh[130], Mervin Mahenthran[130], Emma Carter[130], Thomas Kong[130], Helen Blackman[131], Ben Creagh-Brown[131], Sinead Donlon[131], Natalia Michalak-Glinska[131], Sheila Mtuwa[131], Veronika Pristopan[131], Armorel Salberg[131], Eleanor Smith[131], Sarah Stone[131], Charles Piercy[131], Jerik Verula[131], Dorota Burda[131], Rugia Montaser[131], Lesley Harden[131], Irving Mayangao[131], Cheryl Marriott[131], Paul Bradley[131], Celia Harris[131], Susan Anderson[132], Eleanor Andrews[132], Janine Birch[132], Emma Collins[132], Kate Hammerton[132], Ryan O'Leary[132], Michele Clark[133], Sarah Purvis[133], Russell Barber[134], Claire Hewitt[134], Annette Hilldrith[134], Karen Jackson-Lawrence[134], Sarah Shepardson[134], Maryanne Wills[134], Susan Butler[134], Silvia Tavares[134], Amy Cunningham[134], Julia Hindale[134], Sarwat Arif[134], Sarah Bean[135], Karen Burt[135], Michael Spivey[135], Carrie Demetriou[136], Charlotte Eckbad[136], Sarah Hierons[136], Lucy Howie[136], Sarah Mitchard[136], Lidia Ramos[136], Alfredo Serrano-Ruiz[136], Katie White[136], Fiona Kelly[136], Daniele Cristiano[137], Natalie Dormand[137], Zohreh Farzad[137], Mahitha Gummadi[137], Kamal Liyanage[137], Brijesh Patel[137], Sara Salmi[137], Geraldine Sloane[137], Vicky Thwaites[137], Mathew Varghese[137], Anelise C Zborowski[137], John Allan[138], Tim Geary[138], Gordon Houston[138], Alistair Meikle[138], Peter O'Brien[138], Miranda Forsey[139], Agilan Kaliappan[139], Anne Nicholson[139], Joanne Riches[139], Mark Vertue[139], Miranda Forsey[139], Agilan Kaliappan[139], Anne Nicholson[139], Joanne Riches[139], Mark Vertue[139], Elizabeth Allan[140], Kate Darlington[140], Ffyon Davies[140], Jack Easton[140], Sumit Kumar[140], Richard Lean[140], Daniel Menzies[140], Richard Pugh[140], Xinyi Qiu[140], Llinos Davies[140], Hannah Williams[140], Jeremy Scanlon[140], Gwyneth Davies[140], Callum Mackay[140], Joannne Lewis[140], Stephanie Rees[140], Metod Oblak[141], Monica Popescu[141], Mini Thankachen[141], Andrew Higham[142], Kerry Simpson[142], Jayne Craig[142], Rosie Baruah[143], Sheila Morris[143], Susie Ferguson[143], Amy Shepherd[143], Luke Stephen Prockter Moore[144], Marcela Paola Vizcaychipi[144], Laura Gomes de Almeida Martins[144], Jaime Carungcong[144], Inthakab Ali Mohamed Ali[145], Karen Beaumont[145], Mark Blunt[145], Zoe Coton[145], Hollie Curgenven[145], Mohamed Elsaadany[145], Kay Fernandes[145], Sameena Mohamed Ally[145], Harini Rangarajan[145], Varun Sarathy[145], Sivarupan Selvanayagam[145], Dave Vedage[145], Matthew White[145], Mandy Gill[146], Paul Paul[146], Valli Ratnam[146], Sarah Shelton[146], Inez Wynter[146], Siobhain Carmody[147], Valerie Joan Page[147], Claire Marie Beith[148], Karen Black[148], Suzanne Clements[148], Alan Morrison[148], Dominic Strachan[148], Margaret Taylor[148], Michelle Clarkson[148], Stuart D'Sylva[148], Kathryn Norman[148], Fiona Auld[149], Joanne Donnachie[149], Ian Edmond[149], Lynn Prentice[149], Nikole Runciman[149], Dario Salutous[149], Lesley Symon[149], Anne Todd[149], Patricia Turner[149], Abigail Short[149], Laura Sweeney[149], Euan Murdoch[149], Dhaneesha Senaratne[149], Michaela Hill[150], Thogulava Kannan[150], Wild Laura[150], Rikki Crawley[151], Abigail Crew[151], Mishell Cunningham[151], Allison Daniels[151], Laura Harrison[151], Susan Hope[151], Ken Inweregbu[151], Sian Jones[151], Nicola Lancaster[151], Jamie Matthews[151], Alice Nicholson[151], Gemma Wray[151], Helen Langton[152], Rachel Prout[152], Malcolm Watters[152], Catherine Novis[152], Anthony Barron[153], Ciara Collins[153], Sundeep Kaul[153], Heather Passmore[153], Claire Prendergast[153], Anna Reed[153], Paula Rogers[153], Rajvinder Shokkar[153], Meriel Woodruff[153], Hayley Middleton[153], Oliver Polgar[153], Claire

Nolan[153], Vicky Thwaites[153], Kanta Mahay[153], Dawn Collier[154], Anil Hormis[154], Victoria Maynard[154], Cheryl Graham[154], Rachel Walker[154], Victoria Maynard[154], Ellen Knights[155], Alicia Price[155], Alice Thomas[155], Chris Thorpe[155], Teresa Behan[156], Caroline Burnett[156], Jonathan Hatton[156], Elaine Heeney[156], Atideb Mitra[156], Maria Newton[156], Rachel Pollard[156], Rachael Stead[156], Vishal Amin[157], Elena Anastasescu[157], Vikram Anumakonda[157], Komala Karthik[157], Rizwana Kausar[157], Karen Reid[157], Jacqueline Smith[157], Janet Imeson-Wood[157], Denise Skinner[158], Jane Gaylard[158], Dee Mullan[158], Julie Newman[158], Denise Skinner[158], Jane Gaylard[158], Dee Mullan[158], Julie Newman[158], Alison Brown[159], Vikki Crickmore[159], Gabor Debreceni[159], Joy Wilkins[159], Liz Nicol[159], Waqas Khaliq[160], Rosie Reece-Anthony[160], Mark Birt[160], Alison Ghosh[161], Emma Williams[161], Louise Allen[162], Eva Beranova[162], Nikki Crisp[162], Joanne Deery[162], Tracy Hazelton[162], Alicia Knight[162], Carly Price[162], Sorrell Tilbey[162], Salah Turki[162], Sharon Turney[162], Joshua Cooper[163], Cheryl Finch[163], Sarah Liderth[163], Alison Quinn[163], Natalia Waddington[163], Tina Coventry[164], Susan Fowler[164], Michael MacMahon[164], Amanda McGregor[164], Anne Cowley[165], Judith Highgate[165], Anne Cowley[165], Judith Highgate[165], Alison Brown[166], Jane Gregory[166], Susan O'Connell[166], Tim Smith[166], Luigi Barberis[166], Shameer Gopal[167], Nichola Harris[167], Victoria Lake[167], Stella Metherell[167], Elizabeth Radford[167], Amelia Daniel[168], Joanne Finn[168], Rajnish Saha[168], Nikki White[168], Amy Easthope[168], Phil Donnison[169], Fiona Trim[169], Beena Eapen[169], Jenny Birch[170], Laura Bough[170], Josie Goodsell[170], Rebecca Tutton[170], Patricia Williams[170], Sarah Williams[170], Barbara Winter-Goodwin[170], Ailstair Nichol[171], Kathy Brickell[171], Michelle Smyth[171], Lorna Murphy[171], Samantha Coetzee[172], Alistair Gales[172], Igor Otahal[172], Meena Raj[172], Craig Sell[172], Paula Hilltout[173], Jayne Evitts[173], Amanda Tyler[173], Joanne Waldron[173], Kate Beesley[174], Sarah Board[174], Agnieszka Kubisz-Pudelko[174], Alison Lewis[174], Jess Perry[174], Lucy Pippard[174], Di Wood[174], Clare Buckley[174], Peter Barry[175], Neil Flint[175], Patel Rekha[175], Dawn Hales[175], Lara Bunni[176], Claire Jennings[176], Monica Latif[176], Rebecca Marshall[176], Gayathri Subramanian[176], Peter J McGuigan[177], Christopher Wasson[177], Stephanie Finn[177], Jackie Green[177], Erin Collins[177], Bernadette King[177], Andy Campbell[178], Sara Smuts[178], Joseph Duffield[178], Oliver Smith[178], Lewis Mallon[178], Watkins Claire[178], Liam Botfield[179], Joanna Butler[179], Catherine Dexter[179], Jo Fletcher[179], Atul Garg[179], Aditya Kuravi[179], Poonam Ranga[179], Emma Virgilio[179], Zakaula Belagodu[180], Bridget Fuller[180], Anca Gherman[180], Olumide Olufuwa[180], Remi Paramsothy[180], Carmel Stuart[180], Naomi Oakley[180], Charlotte Kamundi[180], David Tyl[180], Katy Collins[180], Pedro Silva[180], June Taylor[180], Laura King[180], Charlotte Coates[180], Maria Crowley[180], Phillipa Wakefield[180], Jane Beadle[180], Laura Johnson[180], Janet Sargeant[180], Madeleine Anderson[180], Ailbhe Brady[181], Rebekah Chan[181], Jeff Little[181], Shane McIvor[181], Helena Prady[181], Helen Whittle[181], Bijoy Mathew[181], Ben Attwood[182], Penny Parsons[182], Geraldine Ward[183], Pamela Bremmer[183], West Joe[184], Baird Tracy[184], Ruddy Jim[184], Ellie Davies[185], Lisa Roche[185], Sonia Sathe[185], Catherine Dennis[186], Alastair McGregor[186], Victoria Parris[186], Sinduya Srikaran[186], Anisha Sukha[186], Rachael Campbell[187], Noreen Clarke[187], Jonathan Whiteside[187], Mairi Mascarenhas[187], Avril Donaldson[187], Joanna Matheson[187], Fiona Barrett[187], Marianne O'Hara[187], Laura Okeefe[187], Clare Bradley[187], Christine Eastgate-Jackson[188], Helder Filipe[188], Daniel Martin[188], Amitaa Maharajh[188], Sara Mingo Garcia[188], Glykeria Pakou[188], Mark De Neef[188], Kathy Dent[189], Elizabeth Horsley[189], Muhmmad Nauman Akhtar[189], Sandra Pearson[189], Dorota Potoczna[189], Sue Spencer[189], Melanie Clapham[190], Rosemary Harper[190], Una Poultney[190], Polly Rice[190], Tim Smith[190], Rachel Mutch[190], Luigi Barberis[190], Lisa Armstrong[191], Hayley Bates[191], Emma Dooks[191], Fiona Farquhar[191], Brigid Hairsine[191], Chantal McParland[191], Sophie Packham[191], Rehana Bi[192], Barney Scholefield[192], Lydia Ashton[192], Linsha George[193], Sophie Twiss[193], David Wright[193], Manish Chablani[194], Amy Kirkby[194], Kimberley Netherton[194], Kim Davies[195], Linda O'Brien[195], Zohra Omar[195], Igor Otahal[195], Emma Perkins[195], Tracy Lewis[195], Isobel Sutherland[195], Karen Burns[196], Andrew Higham[196], Dr Ben Chandler[197], Kerry Elliott[197], Janine Mallinson[197], Alison Turnbull[197], Prisca Gondo[198], Bernard Hadebe[198], Abdul Kayani[198], Bridgett Masunda[198], Taya Anderson[199], Dan Hawcutt[199], Laura O'Malley[199], Laura Rad[199], Naomi Rogers[199], Paula Saunderson[199], Kathryn Sian Allison[199], Deborah Afolabi[199], jennifer whitbread[199], Dawn jones[199], Rachael Dore[199], Matthew Halkes[200], Pauline Mercer[200], Lorraine Thornton[200], Joy Dawson[201], Sweyn Garrioch[201], Melanie Tolson[201], Jonathan Aldridge[201], Ritoo Kapoor[202], David Loader[202], Karen Castle[202], Sally humphreys[203], Ruth Tampsett[203], Katherine Mackintosh[204], Amanda Ayers[204], Wendy Harrison[204], Julie North[204], Suzanne Allibone[205], Roman Genetu[205], Vidya Kasipandian[205], Amit Patel[205], Ainhi Mac[205], Anthony Murphy[205], Parisa Mahjoob[205], Roonak Nazari[205], Lucy Worsley[205], Andrew Fagan[205], Thomas Bemand[206], Ethel Black[206], Arnold Dela Rosa[206], Ryan Howle[206], Shaman Jhanji[206], Ravishankar Rao Baikady[206], Kate Colette Tatham[206], Benjamin Thomas[206], Dina Bell[207], Rosalind Boyle[207], Katie Douglas[207], Lynn Glass[207],

Emma Lee[207], Liz Lennon[207], Austin Rattray[207], Abigail Taylor[208], Rachel Anne Hughes[208], Helen Thomas[208], Alun Rees[208], Michaela Duskova[208], Janet Phipps[208], Suzanne Brooks[208], Michelle Edwards[208], Victoria Parris[209], Sheena Quaid[209], Ekaterina Watson[209], Adam Brayne[210], Emma Fisher[210], Jane Hunt[210], Peter Jackson[210], Duncan Kaye[210], Nicholas Love[210], Juliet Parkin[210], Victoria Tuckey[210], Lynne Van Koutrik[210], Sasha Carter[210], Benedict Andrew[210], Louise Findlay[210], Katie Adams[210], Jen Service[211], Alison Williams[211], Claire Cheyne[211], Anne Saunderson[211], Sam Moultrie[211], Miranda Odam[211], Kathryn Hall[212], Isheunesu Mapfunde[212], Charlotte Willis[212], Alex Lyon[212], Chunda Sri-Chandana[213], Joslan Scherewode[213], Lorraine Stephenson[213], Sarah Marsh[213], David Brealey[214], John Hardy[214], Henry Houlden[214], Eleanor Moncur[214], Eamon Raith[214], Ambreen Tariq[214], Arianna Tucci[214], Maria Hobrok[215], Ronda Loosley[215], Heather McGuinness[215], Helen Tench[215], Rebecca Wolf-Roberts[215], Val Irvine[216], Benjamin Shelley[216], Amy Easthope[217], Claire Gorman[217], Abhinav Gupta[217], Elizabeth Timlick[217], Rebecca Brady[217], Colin Begg[4], Barry Milligan[4], Arianna Bellini[218], Jade Bryant[218], Anton Mayer[218], Amy Pickard[218], Nicholas Roe[218], Jason Sowter[218], Alex Howlett[218], Katy Fidler[219], Emma Tagliavini[219], Kevin Donnelly[219].

[2] Roslin Institute, University of Edinburgh, Easter Bush, Edinburgh, EH25 9RG, UK

[3] Intensive Care Unit, Royal Infirmary of Edinburgh, 54 Little France Drive, Edinburgh, EH16 5SA, UK

[4] Royal Hospital for Children, Glasgow, UK

[5] William Harvey Research Institute, Barts and the London School of Medicine and Dentistry, Queen Mary University of London, London EC1M 6BQ, UK

[6] Centre for Tropical Medicine and Global Health, Nuffield Department of Medicine, University of Oxford, Old Road Campus, Roosevelt Drive, Oxford, OX3 7FZ, UK

[7] Wellcome Centre for Human Genetics, University of Oxford, Oxford, UK

[8] Prince of Wales Hospital, Hong Kong, China

[9] Department of Critical Care Medicine, Queen's University and Kingston Health Sciences Centre, Kingston, ON, Canada

[10] Wellcome-Wolfson Institute for Experimental Medicine, Queen's University Belfast, Belfast, Northern Ireland, UK

[11] Department of Intensive Care Medicine, Royal Victoria Hospital, Belfast, Northern Ireland, UK

[12] UCL Centre for Human Health and Performance, London, W1T 7HA, UK

[13] Clinical Research Centre at St Vincent's University Hospital, University College Dublin, Dublin, Ireland

[14] National Heart and Lung Institute, Imperial College London, London, UK

[15] Imperial College Healthcare NHS Trust:London,London,UK

[16] Heart Institute, University of Sao Paulo, Brazil

[17] MRC Human Genetics Unit, Institute of Genetics and Molecular Medicine, University of Edinburgh, Western General Hospital, Crewe Road, Edinburgh, EH4 2XU, UK

[18] Intensive Care National Audit & Research Centre, London, UK

[19] NIHR Health Protection Research Unit for Emerging and Zoonotic Infections, Institute of Infection, Veterinary and Ecological Sciences University of Liverpool, Liverpool, L69 7BE, UK

[20] Respiratory Medicine, Alder Hey Children's Hospital, Institute in The Park, University of Liverpool, Alder Hey Children's Hospital, Liverpool, UK

[21] Department of Intensive Care Medicine, Guy's and St. Thomas NHS Foundation Trust, London, UK

[22] Department of Medicine, University of Cambridge, Cambridge, UK

[23] NIHR Clinical Research Network (CRN), North West London Core Team, 3rd Floor Administrative Block South, Clock Tower, Hammersmith Hospital, Du Cane Road, London W12 0HS

[24] Cambridge University Hospitals NHS Foundation Trust, Hills Road, Cambridge, CB2 0QQ, UK

[25] Edinburgh Clinical Research Facility, Western General Hospital, University of Edinburgh, EH4 2XU, UK

[26] Biostatistics Group, State Key Laboratory of Biocontrol, School of Life Sciences, Sun Yat-sen University, Guangzhou, China

[27] Department of Infectious Diseases, Leiden University Medical Center, Leiden, The Netherlands

[28] Guys and St Thomas' Hospital, London, UK

[29] Barts Health NHS Trust, London, UK

[30] James Cook University Hospital, Middlesbrough, UK

[31] Royal Stoke University Hospital, Staffordshire, UK
[32] North Middlesex University Hospital NHS trust, London, UK
[33] north Middlesex University Hospital NHS trust, London, UK
[34] The Royal Liverpool University Hospital, Liverpool, UK
[35] King's College Hospital, London, UK
[36] Charing Cross Hospital, St Mary's Hospital and Hammersmith Hospital, London, UK
[37] Nottingham University Hospital, Nottingham, UK
[38] John Radcliffe Hospital, Oxford, UK
[39] Kingston Hospital, Surrey, UK
[40] kingston Hospital, Surrey, UK
[41] Royal Infirmary of Edinburgh, Edinburgh, UK
[42] Queen Alexandra Hospital, Portsmouth, UK
[43] Morriston Hospital, Swansea, UK
[44] Addenbrooke's Hospital, Cambridge, UK
[45] BHRUT (Barking Havering) - Queens Hospital and King George Hospital, Essex, UK
[46] Royal Sussex County Hospital, Brighton, UK
[47] Queen Elizabeth Hospital, Birmingham, UK
[48] St George's Hospital, London, UK
[49] Stepping Hill Hospital, Stockport, UK
[50] Countess of Chester Hospital, Chester, UK
[51] Royal Blackburn Teaching Hospital, Blackburn, UK
[52] The Tunbridge Wells Hospital and Maidstone Hospital, Kent, UK
[53] Royal Gwent Hospital, Newport, UK
[54] Pinderfields General Hospital, Wakefield, UK
[55] Royal Berkshire NHS Foundation Trust, Berkshire, UK
[56] Broomfield Hospital, Chelmsford, UK
[57] Northumbria Healthcare NHS Foundation Trust, North Shields, UK
[58] Whiston Hospital, Prescot, UK
[59] Croydon University Hospital, Croydon, UK
[60] York Hospital, York, UK
[61] Heartlands Hospital, Birmingham, UK
[62] Ashford and St Peter's Hospital, Surrey, UK
[63] Barnet Hospital, London, UK
[64] East Surrey Hospital, Redhill, UK
[65] Ninewells Hospital, Dundee, UK
[66] Worthing Hospital, Worthing, UK and St Richard's Hospital, Chichester, UK
[67] Southampton General Hospital, Southampton, UK
[68] The Alexandra Hospital, Redditch and Worcester Royal Hospital, Worcester, UK
[69] Sandwell General Hospital and City Hospital, Birmingham, UK
[70] Blackpool Victoria Hospital, Blackpool, UK
[71] Royal Glamorgan Hospital, Pontyclun, UK
[72] The Royal Oldham Hospital, Manchester, UK
[73] Glasgow Royal Infirmary, Glasgow, UK
[74] St James's University Hospital and Leeds General Infirmary, Leeds, UK
[75] University Hospital North Durham, Darlington, UK and Darlington Memorial Hospital, Darlington, UK
[76] Fairfield General Hospital, Bury, UK
[77] Wythenshawe Hospital, Manchester, UK
[78] Royal Alexandra Hospital, Paisley, UK
[79] Good Hope Hospital, Birmingham, UK
[80] Tameside General Hospital, Ashton Under Lyne, UK
[81] Royal Derby Hospital, Derby, UK
[82] Medway Maritime Hospital, Gillingham, UK
[83] Royal Victoria Infirmary, Newcastle Upon Tyne, UK
[84] Poole Hospital, Poole, UK
[85] Bedford Hospital, Bedford, UK

[86] Queens Hospital Burton, Burton-On-Trent, UK
[87] North Manchester General Hospital, Manchester, UK
[88] Aberdeen Royal Infirmary, Aberdeen, UK
[89] Derriford Hospital, Plymouth, UK
[90] Manchester Royal Infirmary, Manchester, UK
[91] Salford Royal Hospital, Manchester, UK
[92] William Harvey Hospital, Ashford, UK
[93] Queen Elizabeth University Hospital, Glasgow, UK
[94] Bradford Royal Infirmary, Bradford, UK
[95] Bristol Royal Infirmary, Bristol, UK
[96] Norfolk and Norwich University hospital (NNUH), Norwich, UK
[97] Queen Elizabeth Hospital Gateshead, Gateshead, UK
[98] Sunderland Royal Hospital, Sunderland, UK
[99] Aintree University Hospital, Liverpool, UK
[100] Hull Royal Infirmary, Hull, UK
[101] Hull Royal Infirmary, Hull, Uk
[102] University College Hospital, London, UK
[103] Royal Devon and Exeter Hospital, Exeter, UK
[104] The Royal Papworth Hospital, Cambridge, UK
[105] Ipswich Hospital, Ipswich, UK
[106] Southmead Hospital, Bristol, UK
[107] Milton Keynes University Hospital, Milton Keynes, UK
[108] Royal Hampshire County Hospital, Hampshire, UK
[109] Queen Elizabeth Hospital, Woolwich, London, UK
[110] Great Ormond St Hospital and UCL Great Ormond St Institute of Child Health NIHR Biomedical Research Centre, London, UK
[111] Stoke Mandeville Hospital, Buckinghamshire, UK
[112] University Hospital of Wales, Cardiff, UK
[113] Basingstoke and North Hampshire Hospital, Basingstoke, UK
[114] Arrowe Park Hospital, Wirral, UK
[115] Chesterfield Royal Hospital Foundation Trust, Chesterfield, UK
[116] Musgrove Park Hospital, Taunton, UK
[117] Peterborough City Hospital, Peterborough, UK and Hinchingbrooke Hospital, Huntingdon, UK
[118] Royal Hallamshire Hospital and Northern General Hospital, Sheffield, UK
[119] Dumfries and Galloway Royal Infirmary, Dumfries, UK
[120] Royal Bolton Hospital, Bolton, UK
[121] Lister Hospital, Stevenage, UK
[122] Craigavon Area Hospital, County Armagh, NI
[123] Southport and Formby District General Hospital, Ormskirk, UK
[124] Calderdale Royal Hospital, Halifax, UK and Huddersfield Royal Infirmary, Huddersfield, UK
[125] Prince Charles Hospital, Merthyr Tydfil, UK
[126] Royal Bournemouth Hospital, Bournemouth, UK
[127] Royal Preston Hospital, Preston, UK
[128] Whittington Hospital, London, UK
[129] Princess Royal Hospital, Telford and Royal Shrewsbury Hospital, Shrewsbury, UK
[130] Macclesfield District General Hospital, Macclesfield, UK
[131] Royal Surrey County Hospital, Guildford, UK
[132] Hereford County Hospital, Hereford, UK
[133] University Hospital of North Tees, Stockton on Tees, UK
[134] Lincoln County Hospital, Lincoln, UK
[135] Royal Cornwall Hospital, Truro, UK
[136] Royal United Hospital, Bath, UK
[137] Royal Brompton Hospital, London, UK
[138] University Hospital Crosshouse, Kilmarnock, UK
[139] Basildon Hospital, Basildon, UK
[140] Glan Clwyd Hospital, Bodelwyddan, UK

[141] West Middlesex Hospital, Isleworth, UK
[142] Royal Lancaster Infirmary, Lancaster, UK
[143] Western General Hospital, Edinburgh, UK
[144] Chelsea & Westminster NHS Foundation Trust, London, UK
[145] The Queen Elizabeth Hospital, King's Lynn, UK
[146] King's Mill Hospital, Nottingham, UK
[147] Watford General Hospital, Watford, UK
[148] University Hospital Wishaw, Wishaw, UK
[149] Forth Valley Royal Hospital, Falkirk, UK
[150] George Eliot Hospital NHS Trust, Nuneaton, UK
[151] Barnsley Hospital, Barnsley, UK
[152] The Great Western Hospital, Swindon, UK
[153] Harefield Hospital, London, UK
[154] Rotherham General Hospital, Rotherham, UK
[155] Ysbyty Gwynedd, Bangor, UK
[156] Diana Princess of Wales Hospital, Grimsby, UK
[157] Russell's Hall Hospital, Dudley, UK
[158] Princess Royal Hospital, Haywards Heath, UK
[159] St Mary's Hospital, Newport, UK
[160] University Hospital Lewisham, London, UK
[161] Colchester General Hospital, Colchester, UK
[162] Queen Elizabeth the Queen Mother Hospital, Margate, UK
[163] Royal Albert Edward Infirmary, Wigan, UK
[164] Victoria Hospital, Kirkcaldy, UK
[165] Eastbourne District General Hospital, East Sussex, UK and Conquest Hospital, East Sussex, UK
[166] Cumberland Infirmary, Carlisle, UK
[167] New Cross Hospital, Wolverhampton, UK
[168] The Princess Alexandra Hospital, Harlow, UK
[169] Salisbury District Hospital, Salisbury, UK
[170] Dorset County Hospital, Dorchester, UK
[171] University College Dublin, St Vincent's University Hospital, Dublin, Ireland
[172] Glangwili General Hospital, Camarthen, UK
[173] Gloucestershire Royal Hospital, Gloucester, UK
[174] Yeovil Hospital, Yeovil, UK
[175] Leicester Royal Infirmary, Leicester, UK
[176] Royal Manchester Children's Hospital, Manchester, UK
[177] Royal Victoria Hospital, Belfast, NI
[178] Wrexham Maelor Hospital, Wrexham, Wales
[179] Walsall Manor Hospital, Walsall, UK
[180] Darent Valley Hospital, Dartford, UK
[181] Warrington General Hospital, Warrington, UK
[182] Warwick Hospital, Warwick, UK
[183] University Hospitals Coventry & Warwickshire NHS Trust, Coventry, UK
[184] University Hospital Monklands, Airdrie, UK
[185] Princess of Wales Hospital, Llantrisant, UK
[186] Northwick Park Hospital, London, UK
[187] Raigmore Hospital, Inverness, UK
[188] Royal Free Hospital, London, UK
[189] Scunthorpe General Hospital, Scunthorpe, UK
[190] West Cumberland Hospital, Whitehaven, UK
[191] Airedale General Hospital, Keighley, UK
[192] Birmingham Children's Hospital, Birmingham, UK
[193] Liverpool Heart and Chest Hospital, Liverpool, UK
[194] Pilgrim Hospital, Lincoln, UK
[195] Prince Philip Hospital, Lianelli, UK
[196] Furness General Hospital, Barrow-in-Furness, UK

[197] Scarborough General Hospital, Scarborough, UK
[198] Southend University Hospital, Westcliff-on-Sea, UK
[199] Alder Hey Children's Hospital, Liverpool, UK
[200] Torbay Hospital, Torquay, UK
[201] Borders General Hospital, Melrose, UK
[202] Kent & Canterbury Hospital, Canterbury, UK
[203] West Suffolk Hospital, Bury St Edmunds, UK
[204] James Paget University Hospital NHS Trust, Great Yarmouth, UK
[205] The Christie NHS Foundation Trust, Manchester, UK
[206] The Royal Marsden Hospital, London, UK
[207] University Hospital Hairmyres, East Kilbride, UK
[208] Withybush General Hospital, Pembrokeshire, Wales
[209] Ealing Hospital, Southall, UK
[210] North Devon District Hospital, Barnstaple, UK
[211] St John's Hospital Livingston, Livingston, UK
[212] Northampton General Hospital NHS Trust, Northampton, UK
[213] Harrogate and District NHS Foundation Trust, Harrogate, UK
[214] National Hospital for Neurology and Neurosurgery, London, UK
[215] Bronglais General Hospital, Aberystwyth, UK
[216] Golden Jubilee National Hospital, Clydebank, UK
[217] Homerton University Hospital Foundation NHS Trust, London UK
[218] Sheffield Children's Hospital, Sheffield, UK
[219] The Royal Alexandra Children's Hospital, Brighton, UK

#### 23andMe Investigators

Janie F. Shelton[1], Anjali J. Shastri[1], Chelsea Ye[1], Catherine H. Weldon[1], Teresa Filshtein-Sonmez[1], Daniella Coker[1], Antony Symons[1], Jorge Esparza-Gordillo[2], Stella Aslibekyan[1], Adam Auton[1].
[1] 23andMe Inc., 223 N Mathilda Ave, Sunnyvale, CA 94086
[2] Human genetics - R&D, GSK Medicines Research Centre, Target Sciences-R&D, Stevenage, UK

### References

[1] Kousathanas, A. *et al.* Whole genome sequencing reveals host factors underlying critical Covid-19. *Nature* 1–10 (2022).

[2] Pairo-Castineira, E. *et al.* Genetic mechanisms of critical illness in covid-19. *Nature* **591**, 92–98 (2021).

[3] Pereira, A. C. *et al.* Genetic risk factors and covid-19 severity in brazil: results from bracovid study. *medRxiv* (2021). URL https://www.medrxiv.org/content/early/2021/10/07/2021.10.06.21264631.

[4] Carracedo, Á. & on Covid-19 (scourge), S. C. t. U. R. o. h. G. A genome-wide association study of COVID-19 related hospitalization in Spain reveals genetic disparities among sexes (2021).

[5] COVID-19 Host Genetics Initiative. Mapping the human genetic architecture of COVID-19. *Nature* (2021). URL https://doi.org/10.1038/s41586-021-03767-x.

[6] Shelton, J. F. *et al.* Trans-ethnic analysis reveals genetic and non-genetic associations with covid-19 susceptibility and severity. *medRxiv* (2020). URL https://www.medrxiv.org/content/early/2020/09/07/2020.09.04.20188318.

[7] Kousathanas, A. *et al.* Whole genome sequencing identifies multiple loci for critical illness caused by covid-19. *medRxiv* (2021). URL https://www.medrxiv.org/content/early/2021/09/08/2021.09.02.21262965.

[8] Ellinghaus, D. *et al.* Genomewide association study of severe covid-19 with respiratory failure. *The New England journal of medicine* **383**, 1522–1534 (2020).

[9] Initiative, C.-. H. G. & Ganna, A. Mapping the human genetic architecture of COVID-19: An update (2021).

[10] Degenhardt, F. *et al.* New susceptibility loci for severe COVID-19 by detailed GWAS analysis in European populations (2021).
